# Electric and Magnetic Source Imaging in Epilepsy: Anatomical Insights from Realistic Simulations

**DOI:** 10.64898/2026.11.06.26367214

**Authors:** Glykeria Sdoukopoulou, Saeed Jahromi, Ludovica Corona, Hmayag Partamian, Jaehyung Lim, Jawata Afnan, Christophe Grova, Christos Papadelis

**Author notes:** **Corresponding author:**Christos Papadelis, PhD, Assistant Vice President, Neuroscience Research, Cook Children’s Health Care System Director, Pediatric Brain Health and Neurosciences Center, The University of Texas at Arlington Professor of Research in Bioengineering, The University of Texas at Arlington Professor of Pediatrics, Burnett School of Medicine, Texas Christian University Cook Children’s Dodson Specialty Clinics, 1500 Cooper St. Fort Worth, TX 76104.

## Abstract

Neurosurgery is an effective treatment for drug-resistant epilepsy (DRE), but its success depends on the accurate delineation of the epileptogenic focus, approximated noninvasively by electric and magnetic source imaging (ESI/MSI) of high-density electroencephalography (HD-EEG) and magnetoencephalography (MEG). These modalities have distinct sensitivity profiles; consequently, epileptiform activity detected by one modality may remain undetected by the other. However, the implications of these differences for spike detectability and localization accuracy, and the added value of combined electromagnetic source imaging (EMSI), remain insufficiently characterized under clinically realistic conditions. Here, we characterize HD-EEG and MEG sensitivity profiles and compare ESI, MSI, and EMSI, using realistic simulations with known ground truth. We generated HD-EEG and MEG datasets from cortical, insular, and hippocampal sources across multiple orientations, activated using patient-derived intracranial EEG spike waveforms. Spike sensitivity and localization accuracy were evaluated using dipole scanning, dynamic statistical parametric mapping (dSPM), and linearly constrained minimum variance (LCMV). EEG demonstrated higher spike detection rates and sensitivity for radially cortical sources at gyral crowns and sulcal bottoms (*p*<0.001), while MEG detected more spikes from tangential cortical sources along sulcal walls (*p*<0.001). MSI localized tangential sources more accurately than ESI across all methods (*p*<0.001), while dipole scanning outperformed dSPM and LCMV for cortical and insular sources (*p*<0.001). EMSI outperformed ESI (*p*<0.001) but did not improve localization over MSI. These findings suggest that complementary anatomical sensitivities of HD-EEG and MEG may improve epileptogenic source characterization, supporting simultaneous acquisition during the presurgical evaluation of patients with DRE.

**Key points:**

- EEG and MEG exhibit complementary sensitivity profiles, with EEG detecting more spikes from gyral crowns and sulcal bottoms and MEG from sulcal walls.
- MSI provides more focal and precise source localization than ESI, particularly for tangentially oriented sources.
- Dipole-based inverse method achieves higher localization accuracy than experimental dSPM and LCMV.

## 1. Introduction

High-density electroencephalography (EEG) and magnetoencephalography (MEG) are noninvasive electrophysiological techniques widely used to localize interictal spikes in the presurgical evaluation of drug-resistant epilepsy (DRE) [1]. Their distinct sensitivity profiles reflect differences in source orientation, depth, and tissue conductivity [2–5]. EEG detects both radially and tangentially oriented sources, such as those arising from gyri and sulci [3,6,7], but conductivity variations, particularly in the skull, distort scalp potentials and can impair localization if inadequately modeled [7,8]. In contrast, MEG primarily detects tangential sources, such as those originating in the fissural cortex, and has reduced sensitivity to radial and deep generators [9]. Its lower sensitivity to conductivity differences across head tissues may also enable more focal source localization [3,10]. These complementary sensitivities may explain why ∼10% of epileptic activity is detected by only one modality during simultaneous recordings [11,12], including MEG spikes without an EEG counterpart [11–17].

Simultaneous recordings can reveal epileptic activity obscured in either modality [18], with MEG spikes sometimes preceding EEG spikes [19] or escaping detection by standard EEG [20]. Detectability also depends on source orientation, spatial extent, and background activity [21]. These factors are particularly relevant for deep generators, where increased sensor distance, complex cortical folding, and cytoarchitecture can further constrain signal detection and localization. Open-field neuronal configurations favor measurable scalp signals, whereas closed-field configurations promote signal cancellation [22,23]. Despite its depth, the hippocampus has an open-field configuration [24], and MEG can detect hippocampal activity, including in patients with normal MRI and extratemporal epilepsy [25], as well as field patterns associated with hippocampal discharges [26]. MEG has also identified tight clusters of insular dipoles undetected by EEG [27]. Together, these findings motivate systematic assessment of the added value of MEG and combined electromagnetic source imaging (EMSI) for localizing anatomically challenging generators [28–33].

The lack of definitive ground truth in clinical studies limits our understanding of how anatomy and generator biophysics influence spike detectability and localization. Limited clinical adoption of electric source imaging (ESI) and magnetic source imaging (MSI), alongside the largely experimental use of EMSI, further complicates their comparative evaluation. Realistic computational simulations provide a controlled framework for linking epileptic generators to their EEG/MEG signatures [34], allowing systematic evaluation of how source orientation, extent, and depth affect detectability and localization across ESI, MSI, and EMSI. Previous simulation studies, however, have largely examined sensitivity or localization separately, with limited assessment of EMSI or integration into conventional presurgical workflows. Gain-matrix analyses have identified source depth as a major constraint on MEG sensitivity while showing that deep tangential sources can remain detectable in realistic head models [4,35]. Simulations using artificial waveforms and idealized sulcal geometries have also shown that opposing sulcal-wall activity can be reconstructed under low-noise conditions [5]. Other studies demonstrated complementary EEG/MEG sensitivities using synthetic cortical spikes and noise, but did not assess localization accuracy [4,36]. Investigations of deep generators, particularly within the hippocampal complex, have further emphasized the importance of realistic head models and advanced inverse methods [9,22,23,37,38]. However, these approaches have predominantly relied on synthetic signals or gain-matrix analyses rather than clinically realistic spikes, leaving the relationship between spike detectability and localization accuracy insufficiently characterized.

Here, we aim to: (i) relate HD-EEG/MEG sensitivity to ESI, MSI, and EMSI localization accuracy for generators in the gyral and fissural cortex, insula, and hippocampal complex; (ii) assess the MEG’s added value over EEG for spike detection within a standard presurgical protocol, particularly for anatomically challenging generators; and (iii) determine whether EMSI improves localization relative to ESI or MSI alone. We hypothesize that (i) cortical folding influences both sensitivity and localization accuracy; (ii) MEG provides complementary information for detecting and localizing challenging generators; and (iii) EMSI improves localization by the complementary sensitivities of EEG and MEG, with potential implications for surgical planning in DRE.

## 2. Materials and Methods

To construct a realistic simulation framework and enhance the ecological validity of the simulated data, we used individualized MRI-derived head models and interictal spike waveforms extracted from intracranial EEG (iEEG) recordings of patients with DRE (**Fig. 1A**). Cortical, insular, and hippocampal sources were activated across a progressive range of orientations, from radial to tangential (**Fig. 1B**). Paired HD-EEG (256 channels) and MEG (305 gradiometers and magnetometers) sensor-level data were then generated by multiplying the activated source time series by the corresponding forward model (**Fig. 1C**). The resulting simulated recordings were reviewed and annotated for spikes by an experienced epileptologist. Based on these annotations, we performed HD-EEG and MEG sensitivity analysis, followed by source localization via ESI, MSI, and EMSI (**Fig. 1D**).

**Figure 1:**
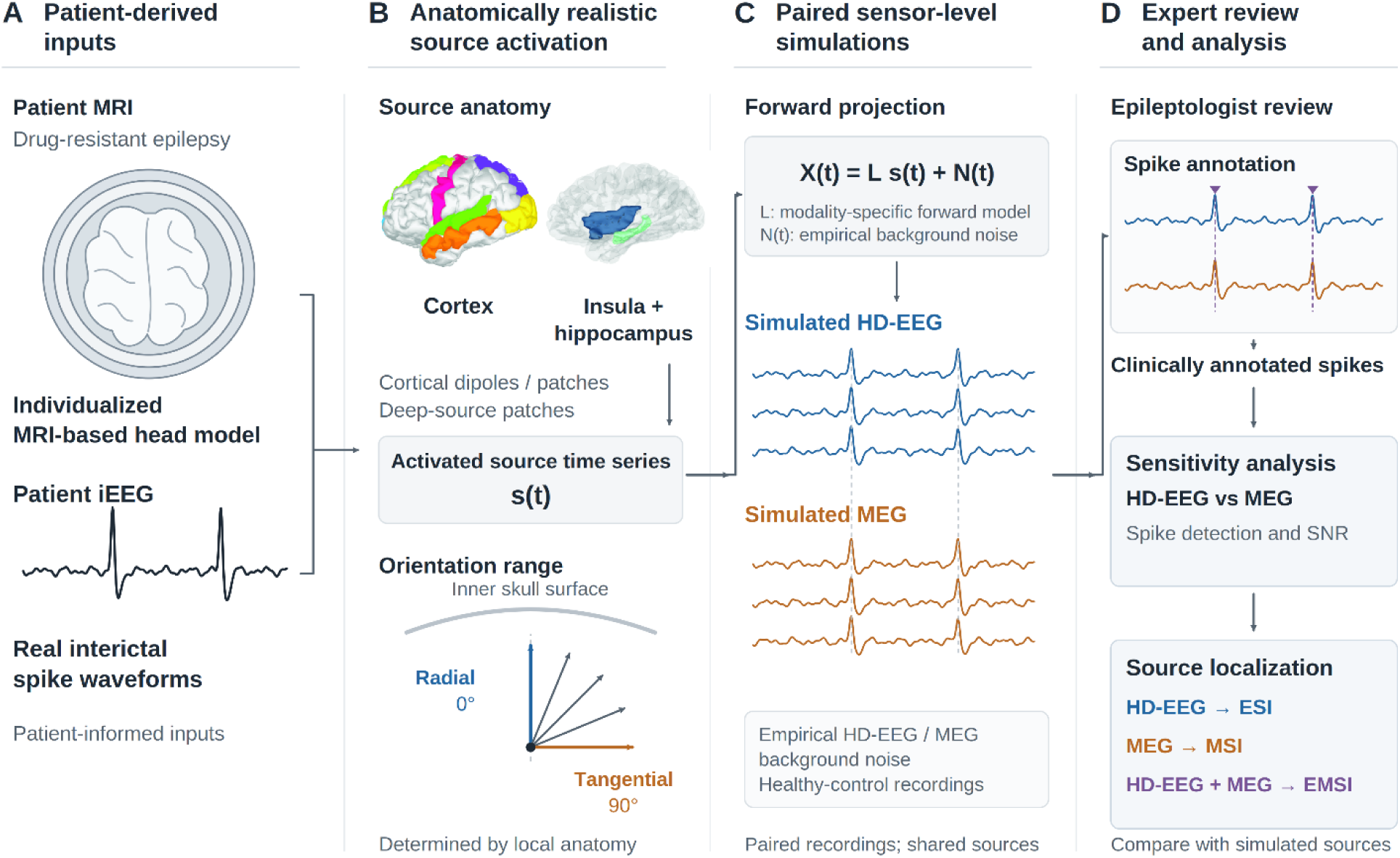
Overview of the simulation pipeline. (**A**) Patient-derived inputs included T1-weighted MRI data from eight children with drug-resistant epilepsy (DRE), used to construct individualized MRI-based head models, and real interictal spike waveforms extracted from patient intracranial EEG (iEEG) recordings. (**B**) Anatomically realistic source activation was modeled across cortical (dipolar and spatially extended), insular, and hippocampal source spaces. Source orientations were determined by local anatomy and ranged progressively from radial (0°) to tangential (90°) relative to the inner skull surface. (**C**) Paired HD-EEG and MEG sensor-level recordings (X(t)) were generated by multiplying the source activation time series (s(t)), by the forward problem (L), and superimposing empirical background noise extracted from healthy-control recordings (N(t)): [X(t) = Ls(t) + N(t)]. (**D**) The resulting simulated recordings were reviewed and annotated for spikes by an experienced epileptologist. The annotated spikes were then used to assess modality-specific HD-EEG and MEG sensitivity through spike detection rates and SNR, followed by source localization using ESI, MSI, and EMSI and comparison with the simulated ground-truth sources.

### 2.1. Realistic source space

#### 2.1.1. MRI

We retrospectively analyzed the T1-weighted MRI data from eight children with DRE (5 males; age range: 3–17 yrs., mean: 12.1 yrs., median: 13 yrs). Images were acquired with a Siemens Skyra 3T scanner at Cook Children’s Medical Center using an MPRAGE sequence (TE = 2.45 ms, TR = 2,200 ms, flip angle = 8°, field of view = 250 mm, 176 slices, matrix size = 256 × 256, acquired resolution = 1.0×1.0×1.0 mm^3^).

#### 2.1.2. Segmentation and regions of interest

Patients’ cortical surfaces were reconstructed using *FreeSurfer* [39]. The original pial surfaces consisted of ∼250,000 vertices and were downsampled to 25,000 vertices for computational efficiency while preserving sufficient spatial resolution to capture cortical folding. The cortical source space was restricted to seven epilepsy-relevant regions: Frontal Pole (FP), Lateral Occipital (LO), Middle Temporal (MT), Primary Motor Cortex (PMC), Superior Frontal (SF), Superior Parietal (SP), and Superior Temporal (ST) (**Fig. 2A**).

**Figure 2:**
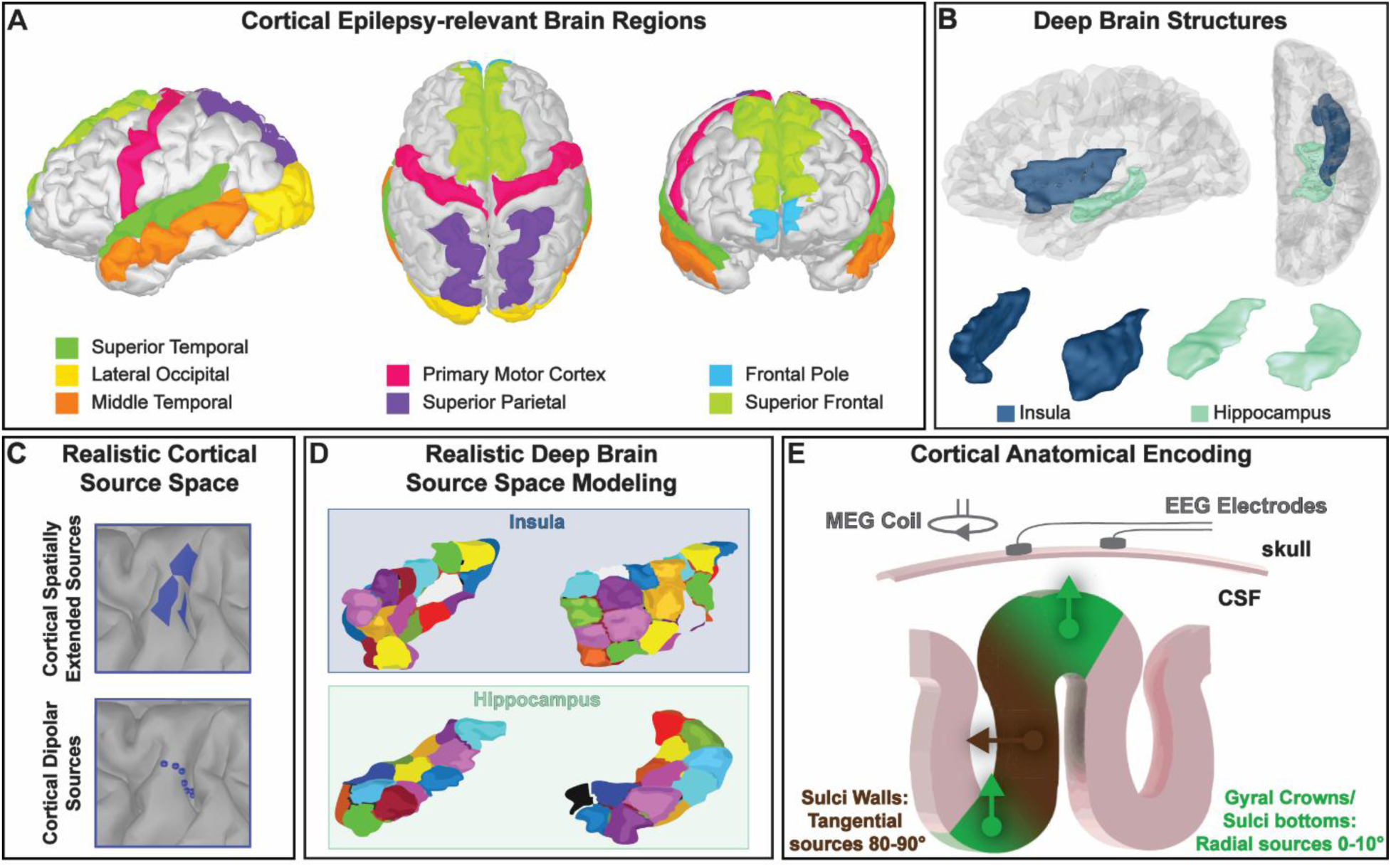
Anatomical encoding and source space definition. (**A**) Experimental design pipeline. (**B**) Cortical source space restricted to seven epilepsy-relevant regions: Frontal Pole (FP), Lateral Occipital (LO), Middle Temporal (MT), Primary Motor Cortex (PMC), Superior Frontal (SF), Superior Parietal (SP), and Superior Temporal (ST). (**C**) Deep structures of interest: insular cortex and hippocampal complex. (**D**) Cortical epileptogenic generators modeled as dipolar (point-like) and spatially extended sources. (**E**) Insular and hippocampal generators modeled as spatially extended sources. (**F**) Anatomical encoding of simulated sources: sources located at the gyral crowns and sulcal bottoms were classified as radial (0°–10°), sources along sulcal walls as tangential (80°–90°), and sources with intermediate orientations as oblique (20°–80°).

Volumetric parcellations of deep brain structures, namely insula and hippocampus, were extracted using the automated anatomical labelling atlas, version 3 (AAL3; **Fig. 2B**) [40]. The insular and hippocampal regions initially contained ∼4,500 and ∼2,500 vertices per hemisphere, respectively, and were downsampled to ∼2,200 and ∼1,200 vertices per hemisphere to reduce computational load while preserving anatomical detail and spatial resolution. Finally, the cortical, insular, and hippocampal surfaces were combined into a unified source space.

#### 2.1.3. Forward model

The forward problem was estimated using *Brainstorm* [41]. For each patient, a realistic head model was constructed from the individuals’ MRIs using *OpenMEEG* [42]. To replicate clinical source localization protocols, a three-layer boundary element model (BEM) comprising the scalp, outer skull, and inner skull surfaces was generated, with conductivities of 1, 0.0165, and 1 S/m, respectively [41]. For computational efficiency, each BEM surface consisted of 2,432 vertices, and the default skull thickness of 4 mm was maintained [41].

#### 2.1.4. Source Modeling

For cortical sources, we modeled both point-like dipolar and spatially extended sources. Point-like sources served as a benchmark, whereas extended sources allowed us to investigate the effects of spatial extent and cortical folding. Within each region of interest (ROI), we selected 24 dipoles, 12 per hemisphere, distributed along the curvature of the cortical mantle (**Fig. 2C**). Spatially extended sources were modeled as activations over cortical patches. To define each patch, we randomly selected a location within the ROI as a seed and progressively expanded the source along the cortical surface, by adding neighboring dipoles. As for the point-like sources, 24 spatially extended sources were generated within each ROI, with 12 per hemisphere (**Fig. 2C**). To avoid cancellation effects introduced by sources spanning across opposing sulcal walls, the average spatial extent was kept at ∼2 cm^2^.

The deep structures of interest, the insular cortex and hippocampal complex, are characterized by open-field neuronal configurations [23]; therefore, their source spaces were defined along the outer surface of each structure, with dipoles oriented orthogonally to the local surface. To model more realistic epileptogenic generators, we focused exclusively on spatially extended sources. We applied k-means clustering [43] to distribute these sources throughout the insular and hippocampal source spaces. Specifically, the vertices of each structure were segmented into *k* spatially coherent, non-overlapping clusters based on Euclidean distance, with each cluster representing one spatially extended source. To obtain a biologically plausible spatial extent comparable to that of the cortical extended sources, we used 40 clusters for the insula (20 per hemisphere) and 30 for the hippocampus (15 per hemisphere; **Fig. 2D**), resulting in an average source extent of ∼2 cm^2^.

#### 2.1.5. Source orientation and anatomical encoding

To quantify the orientation of each dipole, we used the inner skull surface of the BEM model as a reference. For each dipole, we identified the nearest point on the inner skull surface based on Euclidean distance and computed the angle (*∂*) between the dipole’s normal vector 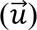 and the normal vector 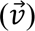 at the corresponding inner skull location using the following formula:

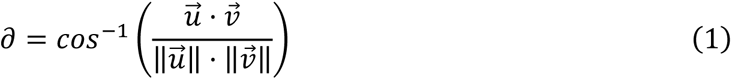

The resulting angle (*∂*) represented the dipole orientation. For cortical dipolar sources, orientation was defined directly by this angle. For cortical and subcortical spatially extended sources, orientation (*∂_mean_*) was defined as the mean of the individual dipole orientation angles (*∂_i_*) comprising each source, as follows:

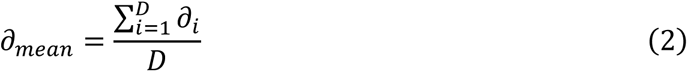

for *i* = 1, …, *D*, where *D* denotes the total number of dipoles within the spatially extended source.

Local cortical geometry, including gyral crowns, sulcal bottoms, sulcal walls, and gyral slopes, determined the orientation of the simulated epileptogenic generators (**Fig. 2E**). Sources located at gyral crowns and sulcal bottoms were classified as radial (0°–10°), those along sulcal walls as tangential (80°–90°), and those along gyral slopes as having both radial and tangential components. For both dipolar and spatially extended cortical sources, each region included at least one fully radial source at a gyral crown or sulcal bottom and one fully tangential source within the fissural cortex. Although the insular cortex and hippocampal complex differ in folding geometry from the cortical mantle, their open-field neuronal configurations similarly allow source orientations spanning the radial-to-tangential spectrum (see *section 2.1.4. Source Modeling*).

### 2.2. Source activation

To ensure that the simulated HD-EEG and MEG sensor-level data resembled epileptic activity, all sources were activated using spike waveforms extracted from iEEG recordings obtained with stereo-EEG (sEEG) electrodes from three patients with DRE (**Fig. 3A**): cortical sources were activated using spikes recorded from a male patient (hippocampal electrode; pathology: tumor), insular sources using spikes recorded from a female patient (insular electrode; pathology: focal cortical dysplasia [FCD]), and hippocampal sources using spikes recorded from a female patient (hippocampal electrode; pathology: FCD). The waveforms were 1.388 s in duration, sampled and processed at 1 kHz, notch-filtered at 60 Hz, and band-pass filtered at 1–70 Hz.

**Figure 3:**
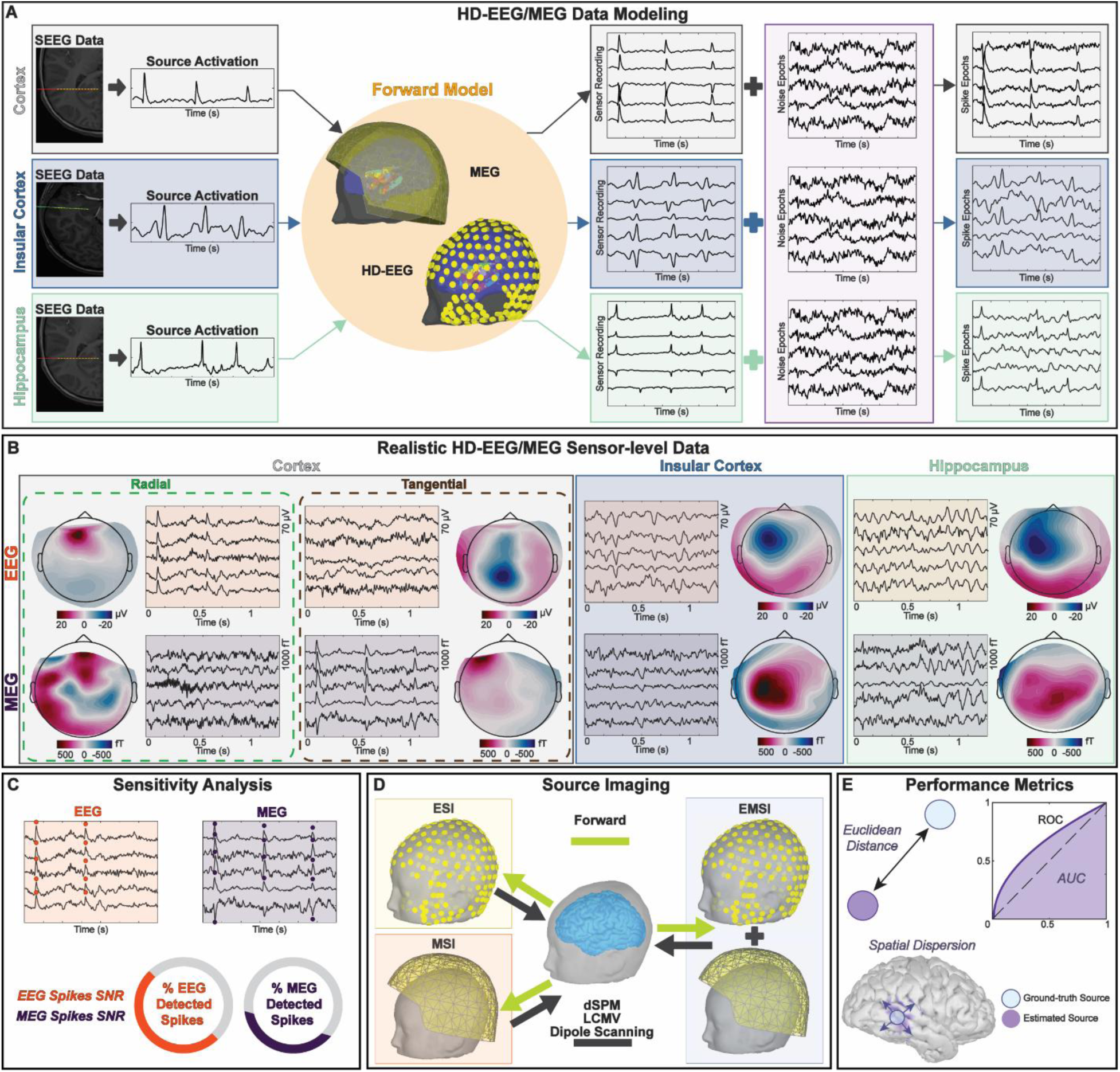
Methodological steps. (**A**) HD-EEG and MEG data modeling process. Cortical dipolar and cortical, insular, and hippocampal spatially extended sources activated using spike waveforms extracted from intracranial EEG (iEEG) recordings, and realistic sensor-level data generated by superimposing empirical background noise. (**B**) Examples of simulated HD-EEG and MEG sensor-level data for cortical radial and tangential sources, depicting the different sensitivity profiles of the two modalities according to local anatomy, as well as for insular and hippocampal sources. (**C**) Sensitivity analysis of detected EEG and MEG spikes based on signal-to-noise ratio (SNR) and detection percentage across source orientations. (**D**) Dipole scanning, dynamic statistical parametric mapping (dSPM), and linearly constrained minimum variance (LCMV) beamforming applied to electric, magnetic, and electromagnetic source imaging (ESI, MSI, and EMSI) for each detected spike. (**E**) Performance metrics including localization error based on Euclidean distance, area under the receiver operating characteristcs curve (AUC), and spatial dispersion, used to evaluate source imaging performance.

Both dipolar and spatially extended cortical sources were activated using the same cortical spike waveform, whereas insular and hippocampal sources were activated using waveforms recorded from the insula and hippocampus, respectively. Each individual dipolar source was assigned a strength of 456 nA.m at the peak of the largest-amplitude spike [36,44]. For spatially extended sources, the peak amplitude per vertex was set to 23 nA.m for cortical sources and 2.5 nA.m for insular and hippocampal sources [36,44].

### 2.3. Realistic HD-EEG and MEG sensor-level modeling

We used a consistent approach to generate realistic HD-EEG and MEG sensor-level data across all source types. Sources within each ROI were sequentially activated using their corresponding spike waveforms (see *section 2.2. Source Activation*), while all remaining sources in the source space were set to zero. The source time series were then multiplied by the forward model to generate simulated sensor-level recordings.

To enhance realism, we added physiological background noise from HD-EEG/MEG recordings of eight healthy controls (six females; age range: 8–13 yrs.; mean age: 10.9 yrs.; median: 11.5 yrs.; **Fig. 2A**). Each simulated patient dataset was paired with a single control subject, from whom an equal number of noise epochs was extracted. This procedure incorporated real physiological and instrumentation noise while preserving the natural sensor-level correlations present in empirical recordings. The final simulated spike epochs were reviewed and annotated by an experienced epileptologist (**Fig. 3B**).

### 2.4. Sensitivity analysis of realistic interictal spikes

To quantitatively assess the sensitivity of HD-EEG and MEG for spike detection, we calculated the signal-to-noise ratio (SNR) of the annotated events as follows:

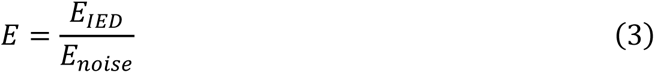

where SNR was defined as the ratio between the energy of the spike (*E_IED_*) and the energy of a 500-ms background noise interval (*E_noise_*). Signal energy was computed in the frequency domain from the power spectral density obtained using the Fourier Transform. SNR was calculated across all channels, and the maximum value was retained for each spike (**Fig. 3C**). To further characterize EEG and MEG detectability, we computed the percentage of detected spikes as a function of source orientation, as well as the overall detection percentage for each modality (**Fig. 3C**).

### 2.5. Source localization of realistic spikes

For source localization, we used the same head models employed to generate the simulated HD-EEG and MEG sensor-level data. Localization accuracy was evaluated using ESI, MSI, and EMSI for HD-EEG, MEG, and combined HD-EEG/MEG data, respectively. To maximize SNR and localize the primary source rather than propagated activity, source imaging was performed at the peak of each spike [1,10]. Noise covariance was modeled using the identity matrix [45,46].

To evaluate the dipole-based approach widely used in clinical practice, we applied dipole scanning with constrained source orientations in *Brainstorm* [41] (**Fig. 3D**). This method evaluates a single dipole at each candidate location within a 3D source grid and quantifies how well each dipole explains the recorded sensor data, with higher values indicating a better fit. The final dipole location and orientation are selected from the grid based on the highest goodness of fit. To assess localization performance using distributed source modeling and beamformer approaches, we additionally applied dynamic statistical parametric mapping (dSPM) [47] and linearly constrained minimum variance (LCMV) beamforming [48], respectively (**Fig. 3D**), as implemented in *Brainstorm*. dSPM estimates distributed source activity by noise-normalizing the reconstructed current at each candidate source location. LCMV uses spatial filters to estimate source activity at each predefined dipole location while minimizing contributions from other locations. For LCMV, data covariance was estimated from spike- and artifact-free data segments. For both dSPM and LCMV, dipole orientations were constrained to be orthogonal to the surface of the modeled structures, including the cortical mantle, insular cortex, and hippocampal complex.

### 2.6. Performance metrics

Localization performance for cortical dipolar sources was assessed using the minimum distance localization error (LE), defined as the Euclidean distance (in mm) between the ground-truth dipolar source and either the estimated dipole location for dipole scanning or the peak of the estimated source map for distributed source models (**Fig. 3E**).

For cortical and subcortical spatially extended sources, the spatial resolution of HD-EEG, MEG, and their combined solution was evaluated using the minimum distance LE and additionally the area under the receiver operating characteristic (ROC) curve (AUC), and the spatial dispersion (SD; **Fig. 3E**) [31,37]. AUC was used as a detection accuracy index to assess how well the distributed source models recovered the spatial extent of the ground-truth source. We used the formulation originally proposed by Grova et al. [49], which corrects for class imbalance resulting from the larger number of inactive than active vertices in the simulations. SD quantified the spatial spread (in mm) of the estimated activity around the ground-truth source and was calculated as the root mean square distance from the ground truth, weighted by the energy of the source localization map. All metrics were computed at the peak of the spikes.

### 2.7. Statistical Analysis

All statistical analyses were performed in MATLAB 2021b (The MathWorks, Inc.). Statistical significance was set at *p*≤0.05. *Wilcoxon rank-sum* tests were used to compare SNR values of detected spikes between EEG and MEG within each source orientation category (radial, tangential, and oblique). The same test was used to compare LE across modalities (ESI, MSI, and EMSI) within each source orientation category. Bonferroni correction was applied to account for multiple comparisons. Extreme LE values considered anatomically implausible for pediatric head dimensions were identified and excluded using the interquartile range method. All performance metrics (LE, AUC, and SD) and SNR values were reported as mean ± standard deviation.

## 3. Results

### 3.1. Sensitivity analysis and anatomical encoding

EEG showed higher SNR for fully radial sources (EEG: 3.91 ± 3.42; MEG: 1.99 ± 1.56; *p*<0.001), whereas MEG showed higher SNR for more tangentially oriented sources (40–60 :° MEG, 4.55 ± 3.49; EEG, 3.49 ± 3.31; 60–80 :° MEG, 5.38 ± 3.78; EEG, 3.13 ± 2.33; 80–90°: MEG, 5.52 ± 3.91; EEG, 2.95 ± 2.22; *p*<0.001; **Fig. 4A**). This orientation dependence was also reflected in spike detection rates. For radial sources, EEG outperformed MEG in both dipolar and spatially extended configurations (EEG: 73.57% and 76.55%; MEG: 11.45% and 4.94%, respectively; **Fig. 4B**), whereas for tangential sources, MEG substantially outperformed EEG (MEG: 92.91% and 75.19%; EEG: 33.33% and 28.32%, respectively; **Fig. 4B**).

**Figure 4:**
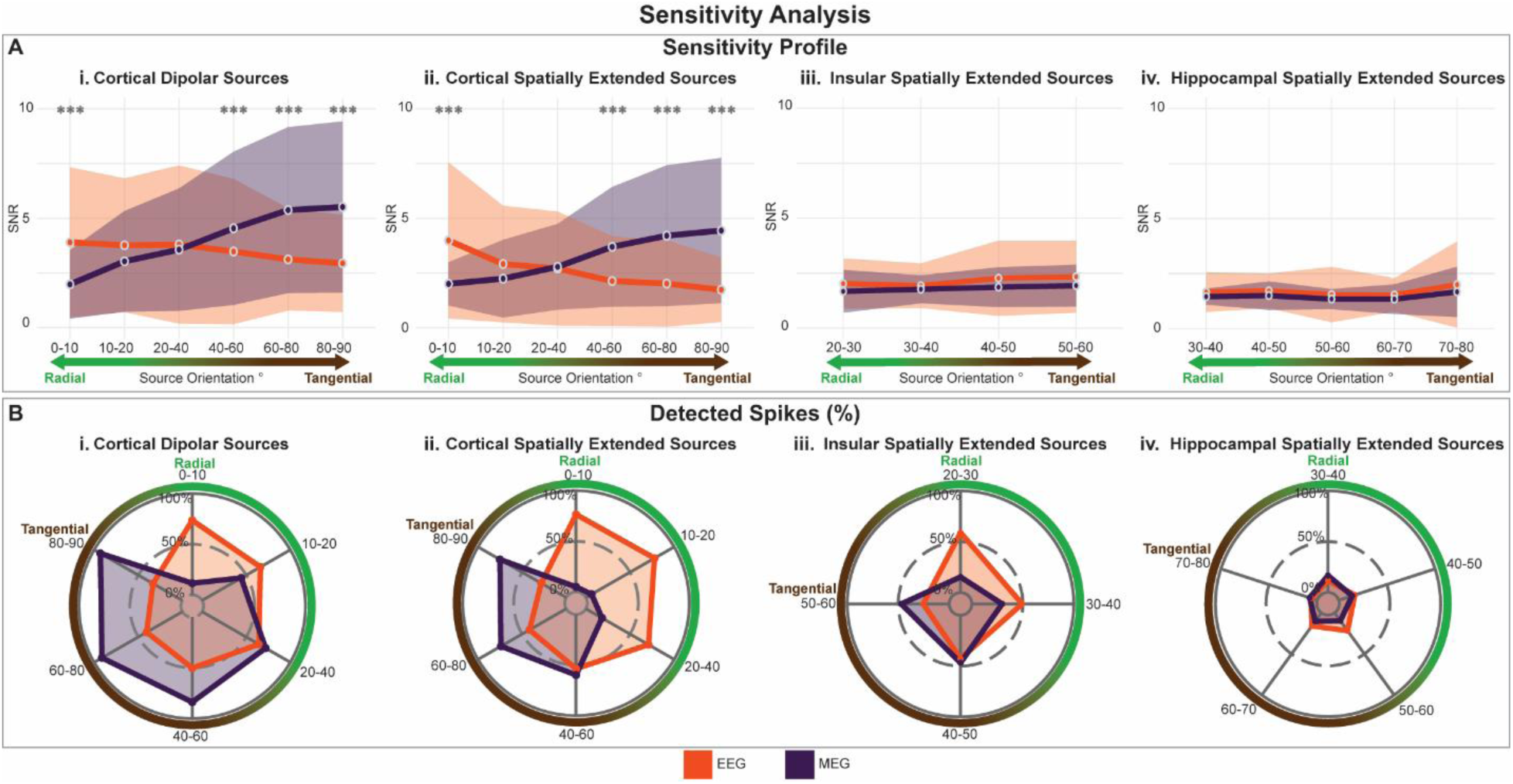
EEG and MEG sensitivity analysis. (**A**) Signal-to-noise ratio (SNR) of detected EEG and MEG spikes for cortical dipolar, cortical spatially extended, insular, and hippocampal sources across source orientations. (**B**) Percentage of detected EEG and MEG spikes for cortical dipolar, cortical spatially extended, insular, and hippocampal sources across source orientations. Significant differences are indicated by asterisks: \*\*\**p*<0.001.

For deep structures, source activity spanned a narrower orientation range and was concentrated within oblique orientations (insula: 20–60°; hippocampal complex: 30–80°). Unlike cortical sources, neither modality showed significant SNR differences across source orientations or a consistent orientation-dependent trend (**Fig. 4A**). Detection patterns, however, differed between structures. In the insula, EEG showed higher detection for more radial sources, whereas MEG showed higher detection for more tangential sources (20–30°: EEG, 58.9%; MEG, 15.4%; 50–60°: EEG, 25.6%; MEG, 47%). In contrast, EEG and MEG detected comparable proportions of hippocampal spikes across orientations (**Fig. 4B**).

### 3.2. Source localization performance

#### 3.2.1. Cortical dipolar sources

**Fig. 5A** shows LE across source orientations and modalities (ESI, MSI, and EMSI) for each inverse method (dipole scanning, dSPM, and LCMV). For dipole scanning, ESI yielded lower LE for radial sources than for tangential sources (8.66 ± 6.43 mm vs. 9.47 ± 5.53 mm), with LE progressively increasing toward tangential orientations. Conversely, MSI localized tangential sources more accurately than radial sources (4.01 ± 3.26 vs. 10.21 ± 8.38 mm), with LE progressively decreasing toward tangential orientations. EMSI showed a mean LE of 10.17 ± 7.9 mm for radial sources, with progressively lower LE toward tangential orientations. No significant differences among modalities were observed for radial sources, whereas for tangential sources, EMSI yielded lower LE than both ESI and MSI (3.37 ± 3.08 vs. 9.47 ± 5.53 and 4.01 ± 3.26 mm, respectively; *p*<0.001, Bonferroni corrected).

**Figure 5:**
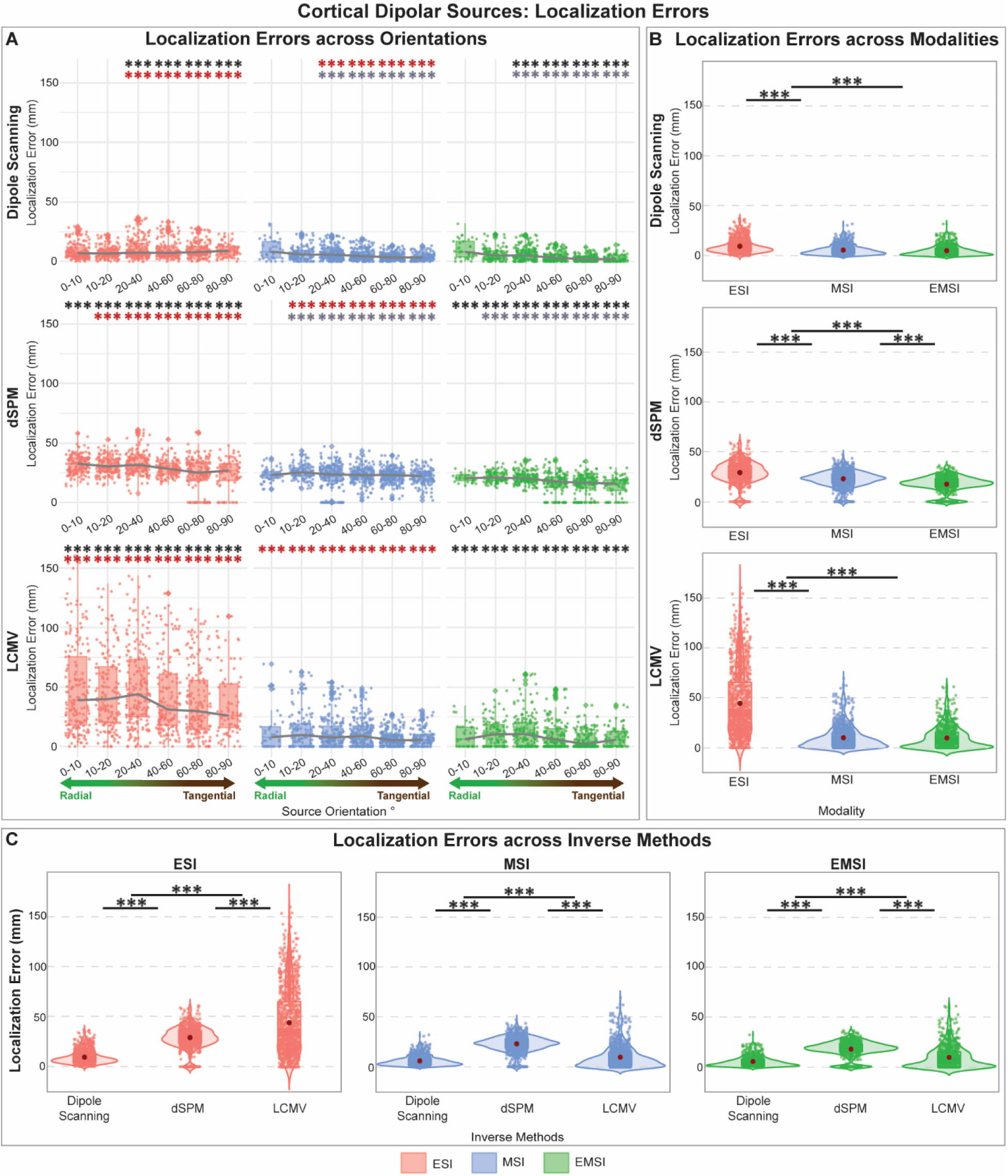
Dipolar sources localization errors across orientations, modalities and inverse methods. (**A**) Localization errors across source orientation and modalities for dipole scanning, dSPM, and LCMV. Black asterisks indicate comparisons between ESI and EMSI, grey asterisks between MSI and EMSI, and red asterisks between ESI and MSI. Grey lines connect the median values across boxplots. Diamonds represent outliers, while dots indicate individual epileptogenic sources. (**B**) Localization errors for dipole scanning, dSPM, and LCMV across modalities. (**C**) Localization errors across inverse methods within each modality (ESI, MSI, and EMSI). In all panels, significant differences are indicated by asterisks: \*\*\**p*<0.001; *p*-values were corrected for multiple comparisons using the Bonferroni method.

For dSPM, EMSI yielded lower LE than ESI for radial sources (20.76 ± 3.5 vs. 32.05 ± 8.04 mm; *p*<0.001, Bonferroni corrected; **Fig. 5A**). LE differ between ESI and MSI (ESI LE: 32.05 ± 8.04 mm; MSI LE: 23.27 ± 5.37 mm; *p* < 0.001, Bonferroni corrected). For tangential sources, EMSI yielded lower LE than both ESI and MSI (13.89 ± 7.94 vs. 24.11 ± 12.95 and 21.71 ± 6.71 mm; *p*<0.001, Bonferroni corrected).

For LCMV, ESI showed substantially higher LE than both dipole scanning and dSPM. For radial sources, EMSI yielded lower LE than both ESI and MSI (9.77 ± 9.36 vs. 49.07 ± 37.76 and 17.12 ± 13.32 mm; *p*<0.001, Bonferroni corrected; **Fig. 5A**). For tangential sources, MSI yielded lower LE than ESI (7.33 ± 7.18 vs. 35.48 ± 28.01 mm; *p*<0.001, Bonferroni corrected; **Fig. 5A**). To compare modalities independently of source orientation, LE was evaluated within each inverse method (**Fig. 5B**). For dipole scanning, MSI yielded lower LE than ESI (4.17 ± 4.85 mm vs. 7.31 ± 6.76 mm; *p*<0.001), with no significant differences between MSI and EMSI. For dSPM, EMSI yielded the lowest LE (18.7 ± 7.85 mm), outperforming both ESI (29.5 ± 10.6 mm) and MSI (23.4 ± 7.61 mm; *p*<0.001, Bonferroni corrected), while MSI also outperformed ESI. For LCMV, MSI again yielded lower LE than ESI (6.55 ± 11.2 mm vs. 35 ± 33.3 mm; *p*<0.001, Bonferroni corrected), with no significant differences between MSI and EMSI.

Comparisons across inverse methods within each modality (**Fig. 5C**) consistently favored dipole scanning. For ESI, dipole scanning yielded lower LE than both dSPM and LCMV (7.31 ± 6.76 vs. 29.5 ± 10.6 and 35 ± 33.3 mm; *p*<0.001). Similarly, for MSI and EMSI, dipole scanning yielded the lowest LE (MSI: 4.17 ± 4.85 mm; EMSI: 3.45 ± 5.01 mm), followed by LCMV (MSI: 6.55 ± 1.2 mm; EMSI: 6.55 ± 11.2 mm) and dSPM (MSI: 23.4 ± 7.61 mm; EMSI: 18.7 ± 7.85 mm; *p*<0.001, Bonferroni corrected).

#### 3.2.2. Cortical spatially extended sources

For dipole scanning, EMSI yielded lower LE than ESI for tangential sources (1.54 ± 1.14 mm vs. 2.07 ± 1.53 mm; *p*<0.001; Bonferroni corrected; **Fig. 6A**), with no significant differences among modalities for radial sources. For dSPM, EMSI yielded lower LE than both ESI and MSI for tangential sources (10.72 ± 5.49 vs. 28.38 ± 14.95 and 14.40 ± 7.07 mm; *p*<0.01, Bonferroni corrected; **Fig. 6A**), whereas no significant differences were observed for radial sources. For source extent reconstruction, ESI yielded higher AUC than MSI for radial sources (0.66 ± 0.14 vs. 0.49 ± 0.11; *p*<0.001, Bonferroni corrected; **Fig. 7A**). For tangential sources, EMSI yielded higher AUC than both ESI and MSI (0.81 ± 0.06 vs. 0.57 ± 0.15 and 0.77 ± 0.06; *p*<0.001, Bonferroni corrected; **Fig. 7A**). Similarly, EMSI and MSI exhibited lower SD than ESI for tangential sources (49.22 ± 9.33 vs. 52.23 ± 9.19 vs. 60.62 ± 9.26 mm; *p*<0.001, Bonferroni corrected; **Fig. 7A**), with no significant differences among modalities for radial sources.

**Figure 6:**
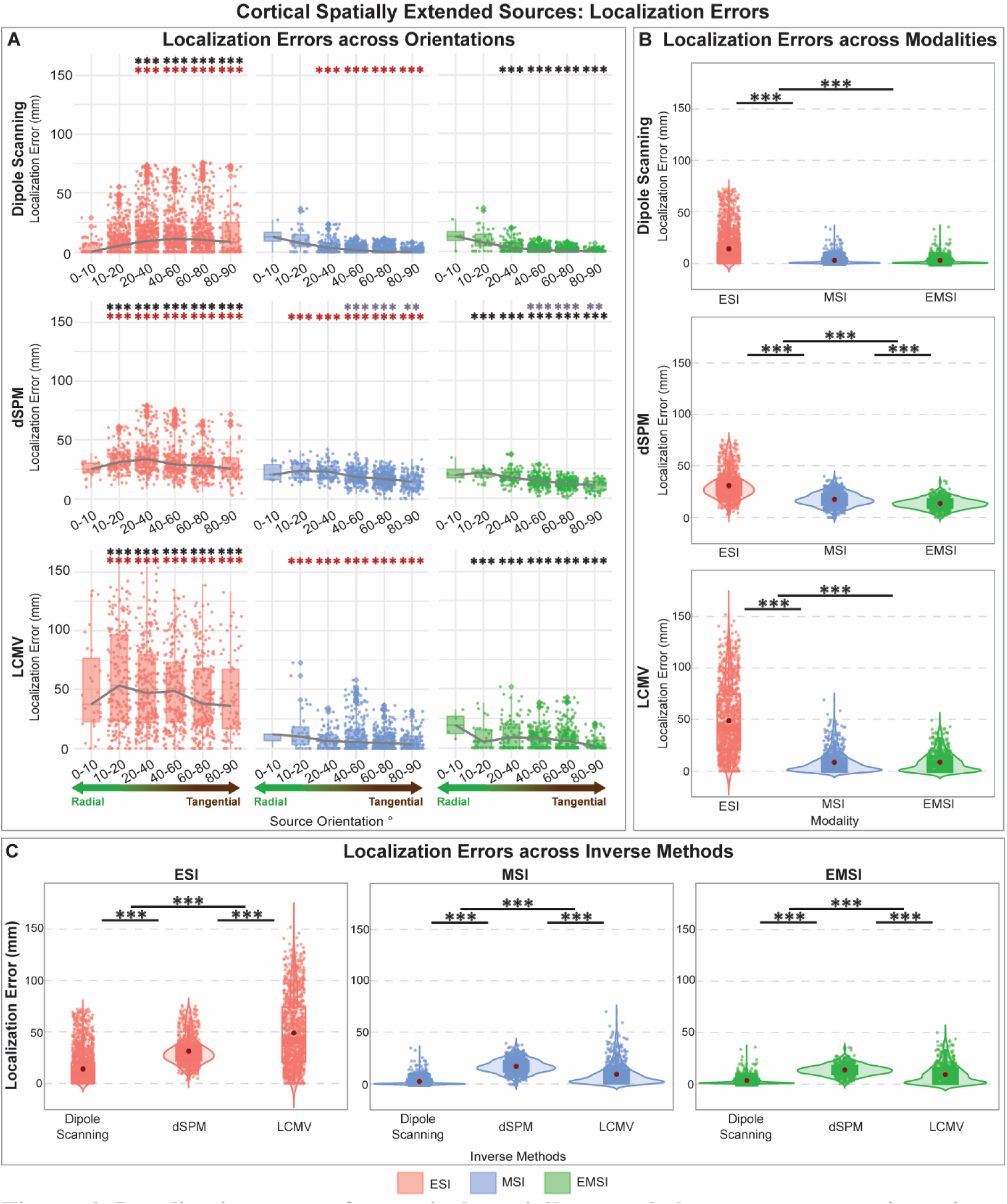
Localization errors for cortical spatially extended sources across orientations, modalities and inverse methods. (**A**) Localization errors across source orientations and modalities for dipole scanning, dSPM, and LCMV. Black asterisks indicate comparisons between ESI and EMSI, grey asterisks between MSI and EMSI, and red asterisks between ESI and MSI. Grey lines connect the medians values across boxplots. Diamonds represent outliers, while dots indicate individual epileptogenic sources. (**B**) Localization error for dipole scanning, dSPM, and LCMV across modalities. (**C**) Localization errors across inverse methods within each modality (ESI, MSI, and EMSI). In all panels, significant differences are indicated by asterisks: \*\**p*<0.01, \*\*\**p*< 0.001; *p*-values were corrected for multiple comparisons using the Bonferroni method.

**Figure 7:**
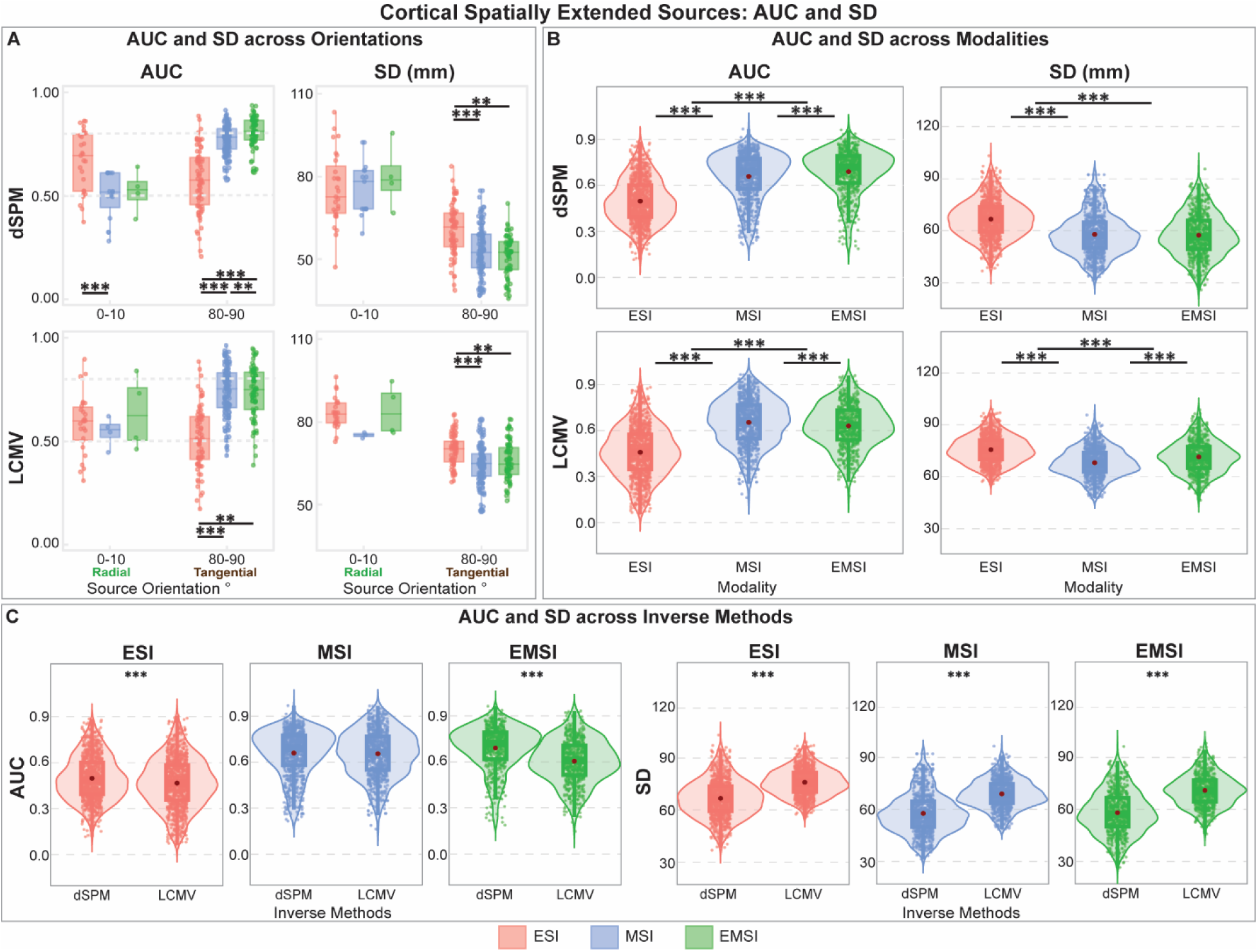
Area under the ROC curve (AUC) and spatial dispersion (SD) for cortical spatially extended sources across orientations, modalities, and inverse methods. (**A**) AUC and SD values for radial and tangential orientations across modalities for dSPM and LCMV. Dashed horizontal lines in the AUC panels at 0.5 and 0.8 denote chance-level detection and a threshold for good detection, respectively. (**B**) AUC and SD for dipole scanning, dSPM, and LCMV across modalities. (**C**) AUC and SD across inverse methods within each modality (ESI, MSI, and EMSI). In all panels, significant differences are indicated by asterisks: \**p*<0.05, \*\**p*<0.01, \*\*\**p*<0.001; *p*-values were corrected for multiple comparisons using the Bonferroni method.

For LCMV, ESI yielded higher LE than both MSI and EMSI across source orientations (*p*<0.001, Bonferroni corrected; **Fig. 6A**), with the largest discrepancy observed for tangential sources (ESI: 43.25 ± 34.34 mm; MSI: 7.70 ± 6.92 mm; EMSI: 5.72 ± 4.17 mm; *p*<0.001, Bonferroni corrected; **Fig. 6A**). For tangential sources, both EMSI and MSI yielded higher AUC than ESI (0.74 ± 0.12 and 0.74 ± 0.12 vs. 0.51 ± 0.15; *p*<0.01, Bonferroni corrected; **Fig. 7A**) and lower SD (65.64 ± 7.03 and 64.65 ± 6.83 vs. 69.57 ± 5.8 mm; *p*<0.001, Bonferroni corrected; **Fig. 7A**). No significant differences among modalities were observed for radial sources.

Regardless of source orientation, dipole scanning yielded higher LE for ESI than both MSI and EMSI (8.7 ± 16.9 vs. 0.87 ± 3.87 and 0.21 ± 3.9 mm; *p*<0.001, Bonferroni corrected; **Fig. 6B**), with no significant differences between MSI and EMSI. For dSPM, ESI similarly yielded higher LE than both MSI and EMSI (30.2 ± 13.8 vs. 17.9 ± 7.01 and 14.1 ± 5.71 mm; *p*<0.001, Bonferroni corrected; **Fig. 6B**), with EMSI also outperforming MSI (*p*<0.001, Bonferroni corrected; **Fig. 6B**). For LCMV, ESI again yielded higher LE than both MSI and EMSI (43.6 ± 36 vs. 5.24 ± 10.9 and 6.65 ± 10.1 mm; *p*<0.001, Bonferroni corrected; **Fig. 6B**), with no significant differences between MSI and EMSI.

Comparisons across inverse methods within each modality showed that, for ESI, dipole scanning yielded the lowest LE (8.7 ± 16.9 mm; *p*<0.001), followed by dSPM (30.2 ± 13.8 mm; *p*<0.001) and LCMV (43.6 ± 36 mm; *p*<0.001, Bonferroni corrected; **Fig. 6C**). ESI dSPM also outperformed LCMV in both AUC and SD (AUC: 0.498 ± 0.139 vs. 0.472 ± 0.158; SD: 66.6 ± 10.8 vs. 75.3 ± 7.8 mm; *p*<0.001; **Fig. 7C**). For MSI, dipole scanning yielded the lowest LE (0.87 ± 3.87 mm; *p*<0.001), followed by LCMV (5.24 ± 10.9 mm; *p*<0.001) and dSPM (17.9 ± 7.01 mm; *p*<0.001; **Fig. 6C**). Although AUC did not differ between dSPM and LCMV, dSPM yielded lower SD (57.2 ± 11.6 vs. 67.8 ± 8.23 mm; *p*<0.001; **Fig. 7C**). For EMSI, dipole scanning similarly yielded the lowest LE (0.21 ± 3.9 mm; *p*<0.001), followed by LCMV (6.65 ± 10.1 mm; *p*<0.001) and dSPM (14.1 ± 5.71 mm; *p*<0.001) and; **Fig. 6C**). For source extent reconstruction, EMSI dSPM outperformed LCMV in both AUC and SD (AUC: 0.725 ± 0.139 vs. 0.644 ± 0.138; SD: 56.5 ± 12.3 vs. 71.3 ± 9.02 mm; *p*<0.001; **Fig. 7C**).

#### 3.2.3. Subcortical spatially extended sources

For the insular cortex, MSI yielded lower LE than ESI for oblique sources across all inverse methods. For sources oriented at 40–50°, LE was lower for MSI than ESI with dipole scanning (15.3 ± 12.5 vs. 22.7 ± 19.2 mm), dSPM (15.1 ± 13.3 vs. 24.9 ± 17.5 mm), and LCMV (13.3 ± 12.2 vs. 42.9 ± 25.6 mm). A similar pattern was observed for sources oriented at 50–60° (dipole scanning: 15.1 ± 11.5 vs. 25.1 ± 22.9 mm; dSPM: 10.5 ± 10.1 vs. 29.9 ± 18.6 mm; LCMV: 12.8 ± 12.4 vs. 44.2 ± 30 mm; *p*<0.001 **Fig. 8A**). Across source orientations, ESI yielded higher LE than MSI and EMSI for all inverse methods (dipole scanning: 21.7 ± 19.1 vs. 15 ± 12.5 and 17.8 ± 15.2 mm; dSPM: 24.2 ± 16.8 vs. 15.5 ± 13.8 and 19.2 ± 24.9 mm; LCMV: 41.5 ± 26.3 vs. 13.7 ± 12.8 and 21.9 ± 20.9 mm; *p*<0.001, Bonferroni corrected; **Fig. 8B**). Across modalities, dipole scanning yielded the lowest LE (*p*<0.001, Bonferroni corrected; **Fig. 8B**). For source extent reconstruction, dSPM yielded lower AUC for ESI than for both MSI and EMSI, whereas LCMV yielded higher AUC for MSI than for both ESI and EMSI (*p*<0.001, Bonferroni corrected; **Fig. 10B**). For dSPM, EMSI showed higher SD than both ESI and MSI, whereas LCMV yielded lower SD for MSI than for both ESI and EMSI (*p*<0.001, Bonferroni corrected; **Fig. 10B**). Across modalities, dSPM yielded higher AUC and lower SD than LCMV (*p*<0.001, Bonferroni corrected; **Fig. 10C**).

**Figure 8:**
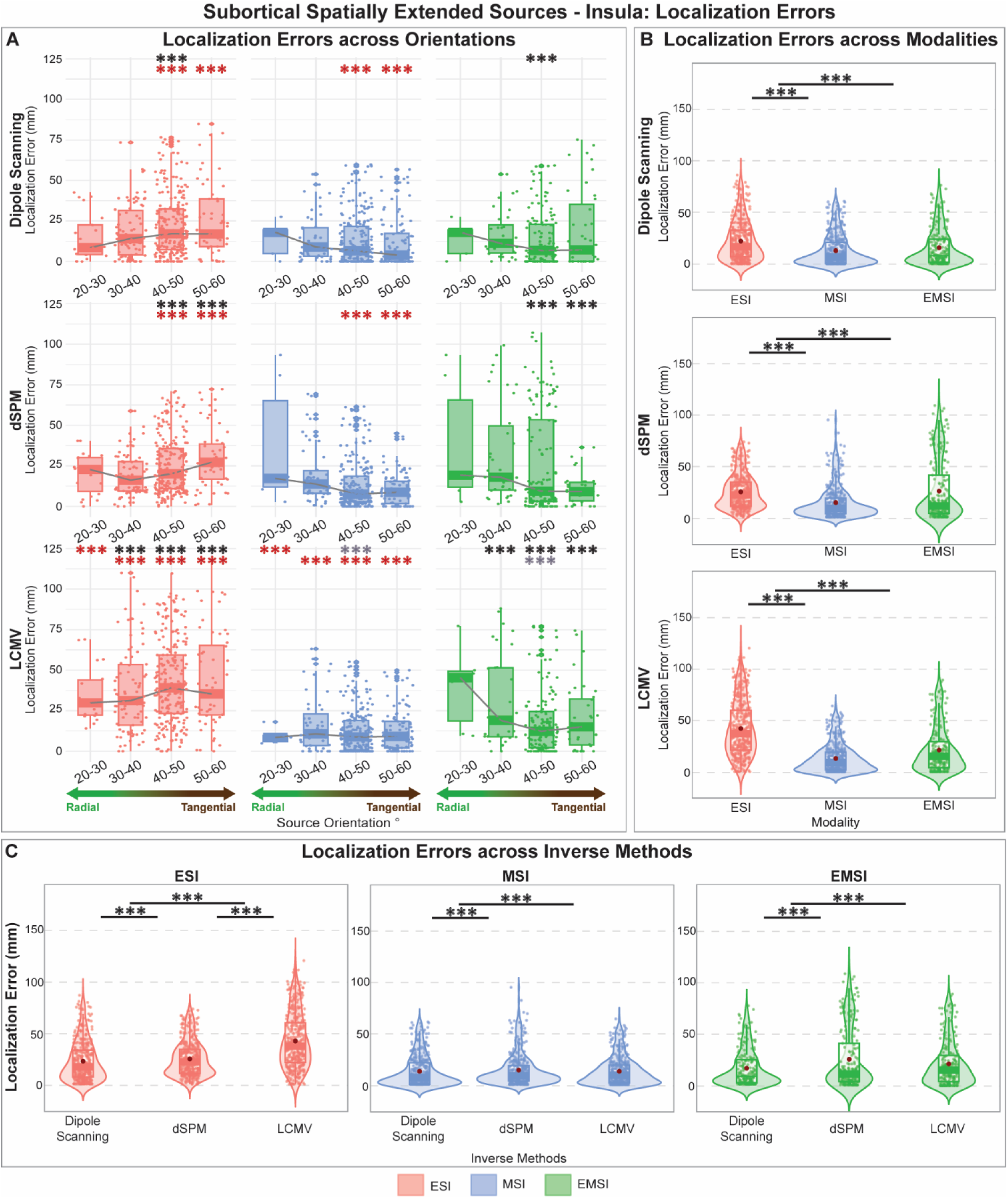
Localization errors for insular spatially extended sources across orientations, modalities and inverse methods. (**A**) Localization errors across source orientation and modality for dipole scanning, dSPM, and LCMV. Black asterisks indicate comparisons between ESI and EMSI, grey asterisks between MSI and EMSI, and red asterisks between ESI and MSI. Grey lines connect the median values across boxplots. Diamonds represent outliers, while dots indicate individual epileptogenic sources. (**B**) Localization errors for dipole scanning, dSPM, and LCMV across modalities. (**C**) Localization errors across inverse methods within each modality (ESI, MSI, and EMSI). In all panels, significant differences are indicated by asterisks: \*\*\**p*<0.001; *p*-values were corrected for multiple comparisons using the Bonferroni method.

For hippocampal complex sources, ESI yielded higher LE than EMSI for dipole scanning and higher LE than both MSI and EMSI for LCMV, regardless of source orientation (*p*<0.001, Bonferroni corrected; **Fig. 9B**). Across modalities, dipole scanning yielded the lowest LE for ESI, outperforming both dSPM and LCMV (*p*<0.001, Bonferroni corrected; **Fig. 9B**). For source extent reconstruction, no significant differences were found between radially and tangentially oriented sources for both dSPM and LCMV AUC and SD (**Fig. 11A**). dSPM yielded lower AUC than LCMV for both MSI and EMSI (*p*<0.001, Bonferroni corrected; **Fig. 11B**). For dSPM, ESI showed lower SD than both MSI and EMSI (*p*<0.001, Bonferroni corrected; **Fig. 11B**). For LCMV, ESI yielded lower AUC than both MSI and EMSI (*p*<0.001, Bonferroni corrected; **Fig. 11B**), but higher SD (*p*<0.001, Bonferroni corrected; **Fig. 11C**).

**Figure 9:**
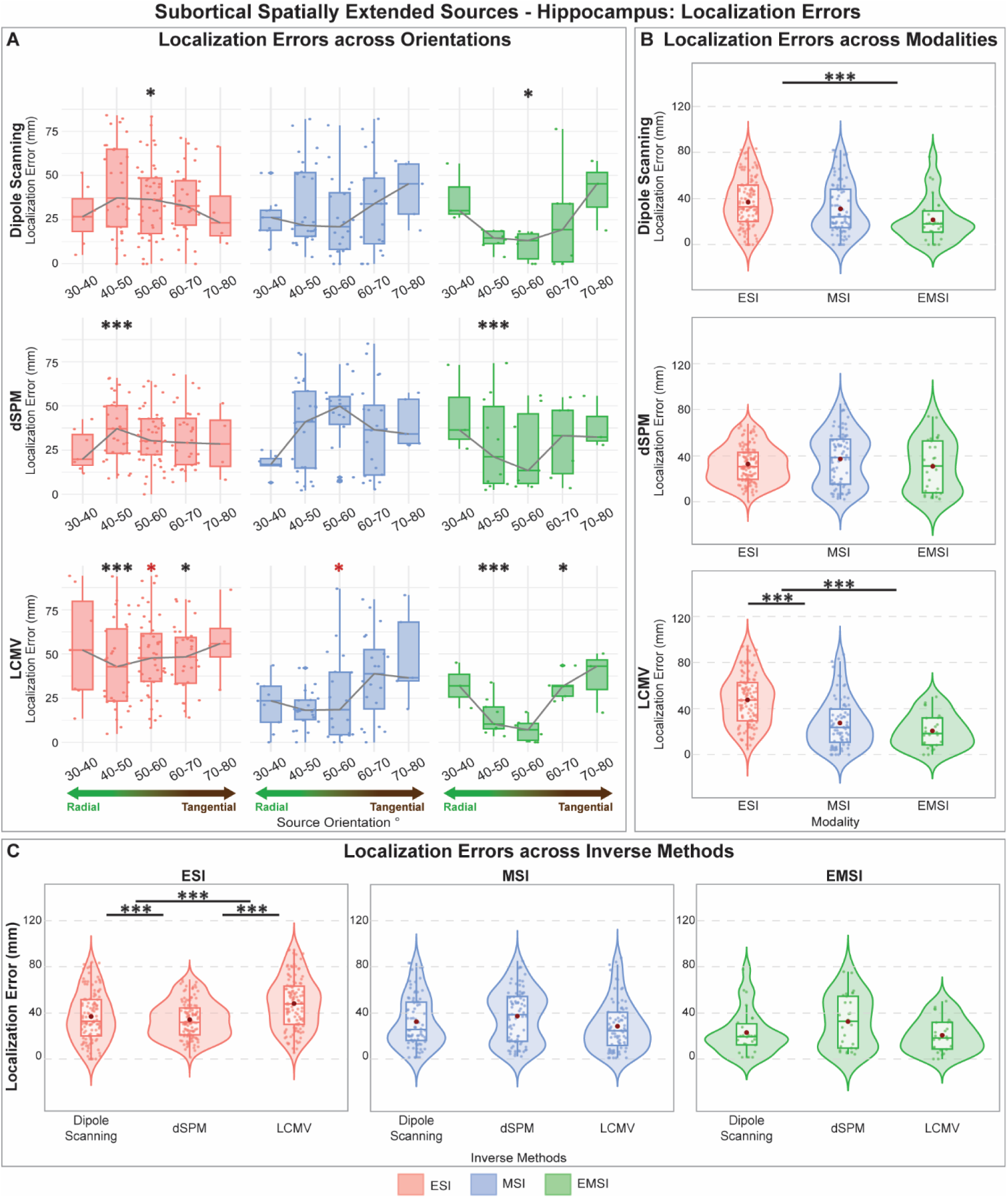
Localization errors for hippocampal spatially extended sources across orientations, modalities and inverse methods. (**A**) Localization errors across source orientations and modalities for dipole scanning, dSPM, and LCMV. Black asterisks indicate comparisons between ESI and EMSI, grey asterisks between MSI and EMSI, and red asterisks between ESI and MSI. Grey lines connect the median values across boxplots. Diamonds represent outliers, while dots indicate individual epileptogenic sources. (**B**) Localization errors for dipole scanning, dSPM, and LCMV across modalities. (**C**) Localization errors across inverse methods within each modality (ESI, MSI, and EMSI). In all panels, significant differences are indicated by asterisks: \**p*<0.05, \*\**p*<0.01, \*\*\**p*<0.001; *p*-values were corrected for multiple comparisons using the Bonferroni method.

**Figure 10:**
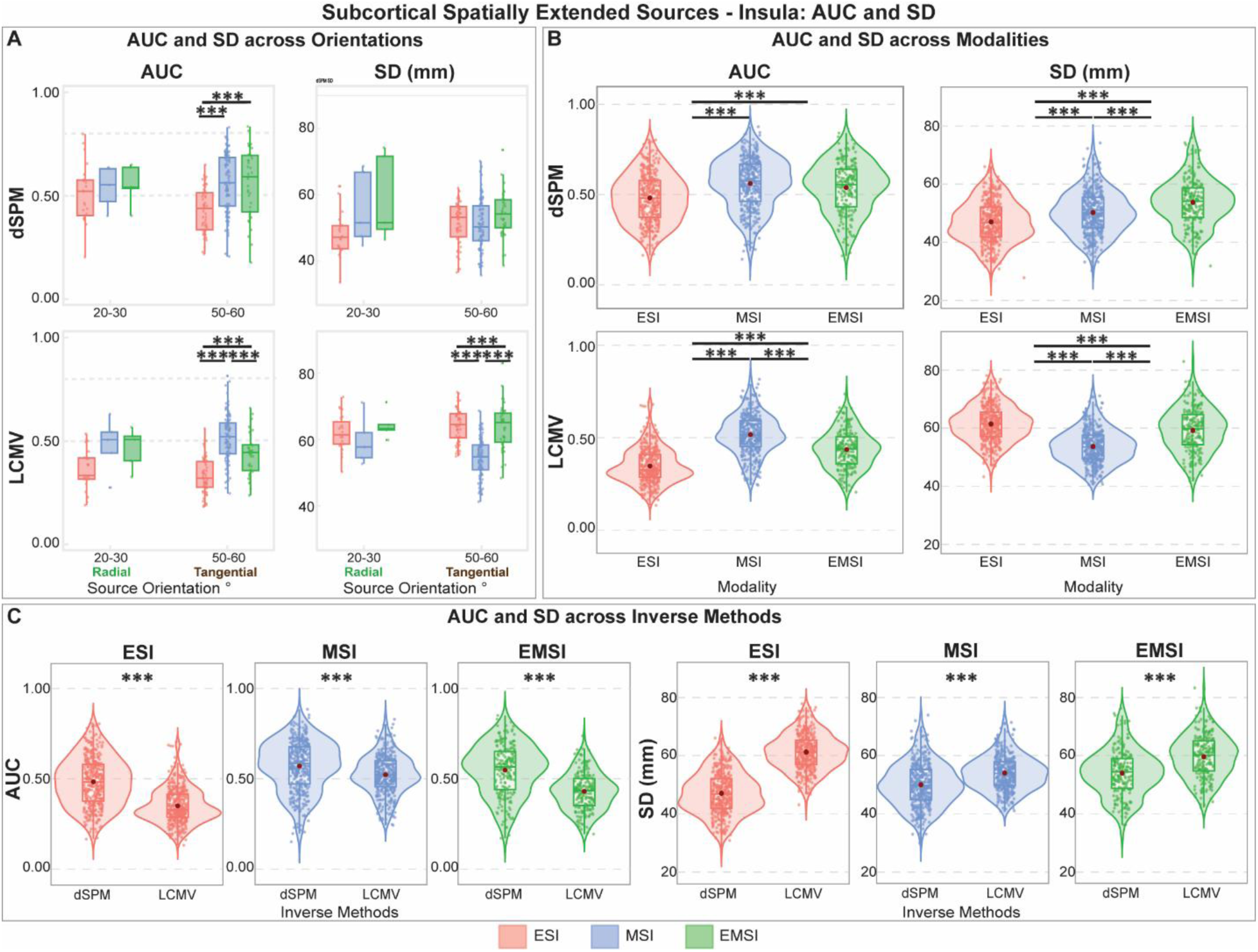
Area under the ROC curve (AUC) and spatial dispersion (SD) for insular spatially extended sources across orientations, modalities, and inverse methods. (**A**) AUC and SD values for radial and tangential source orientations across modalities for dSPM and LCMV. Dashed horizontal lines in the AUC panels at 0.5 and 0.8 denote chance-level detection and a threshold for good detection, respectively. (**B**) AUC and SD for dipole scanning, dSPM, and LCMV across modalities. (**C**) AUC and SD across inverse methods within each modality (ESI, MSI, and EMSI). In all panels, significant differences are indicated by asterisks: \*\**p*<0.01, \*\*\**p*<0.001; *p*-values were corrected for multiple comparisons using the Bonferroni method.

**Figure 11:**
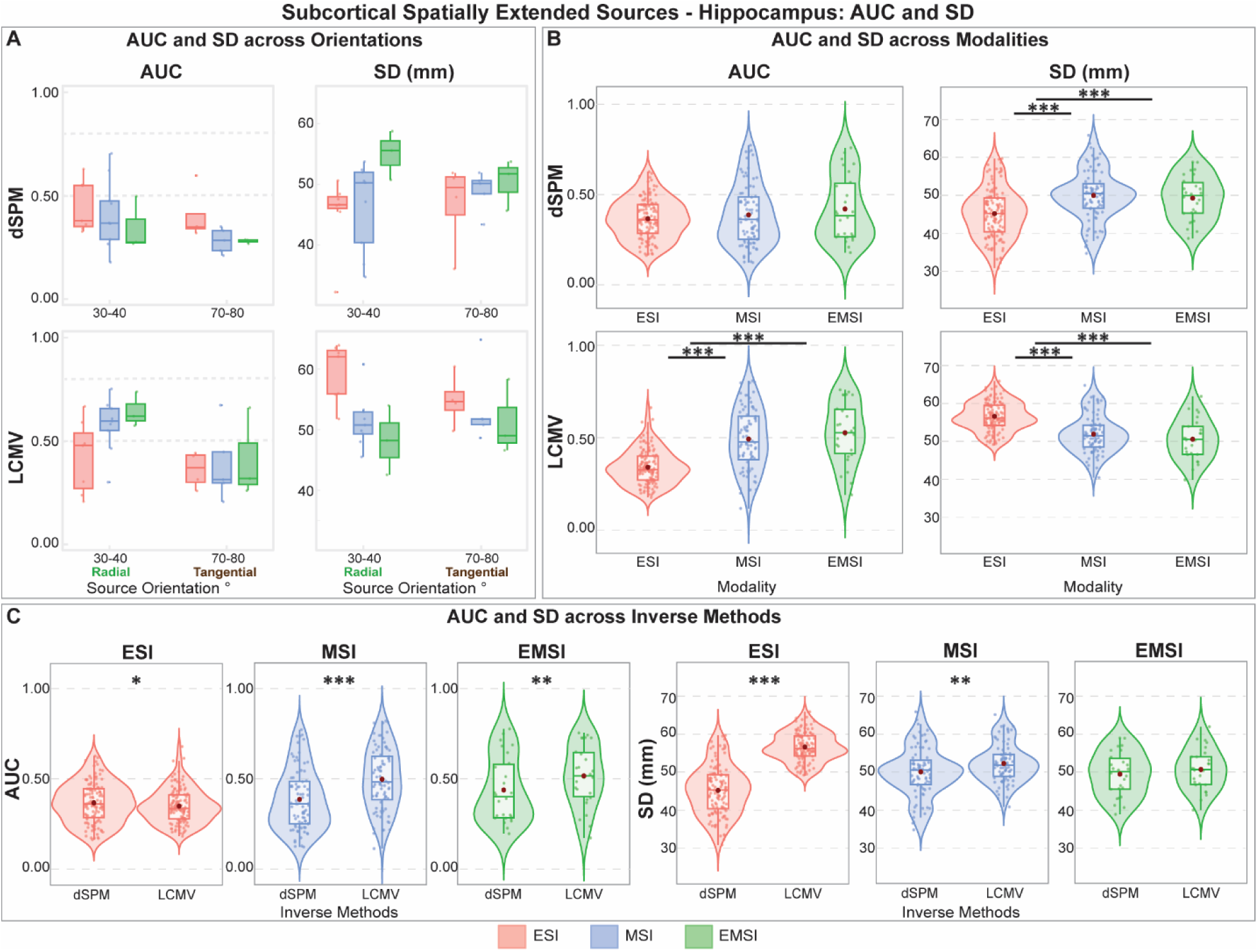
Area under the ROC curve (AUC) and spatial dispersion (SD) for hippocampal spatially extended sources across orientations, modalities, and inverse methods. (**A**) AUC and SD values for radial and tangential source orientations across modalities for dSPM and LCMV. Dashed horizontal lines in the AUC panels at 0.5 and 0.8 denote chance-level detection and a threshold for good detection, respectively. (**B**) AUC and SD for dipole scanning, dSPM, and LCMV across modalities. (**C**) AUC and SD across inverse methods within each modality (ESI, MSI, and EMSI). In all panels, significant differences are indicated by asterisks: \**p*<0.05, \*\**p*<0.01, \*\*\**p*<0.001; *p*-values were corrected for multiple comparisons using the Bonferroni method.

### 3.3. Regional analysis

To characterize regional patterns of EEG/MEG sensitivity and localization accuracy across ESI, MSI, and EMSI, we evaluated SNR and LE values across all source orientations and inverse methods for both dipolar and spatially extended sources (**Fig. 12A**). For dipolar sources, EEG showed higher SNR from sources located at gyral crowns and sulcal bottoms, whereas activity arising from sulcal walls showed lower SNR across brain regions (**Fig. 12B**). In contrast, MEG showed higher SNR for tangential sources (**Fig. 12B**). Across regions, dipole scanning yielded the lowest LE compared with dSPM and LCMV (**Fig. 12C**), while MSI and EMSI showed higher localization accuracy, especially for tangential sources. ESI performance was relatively insensitive to source orientations (**Fig. 12C**).

**Figure 12:**
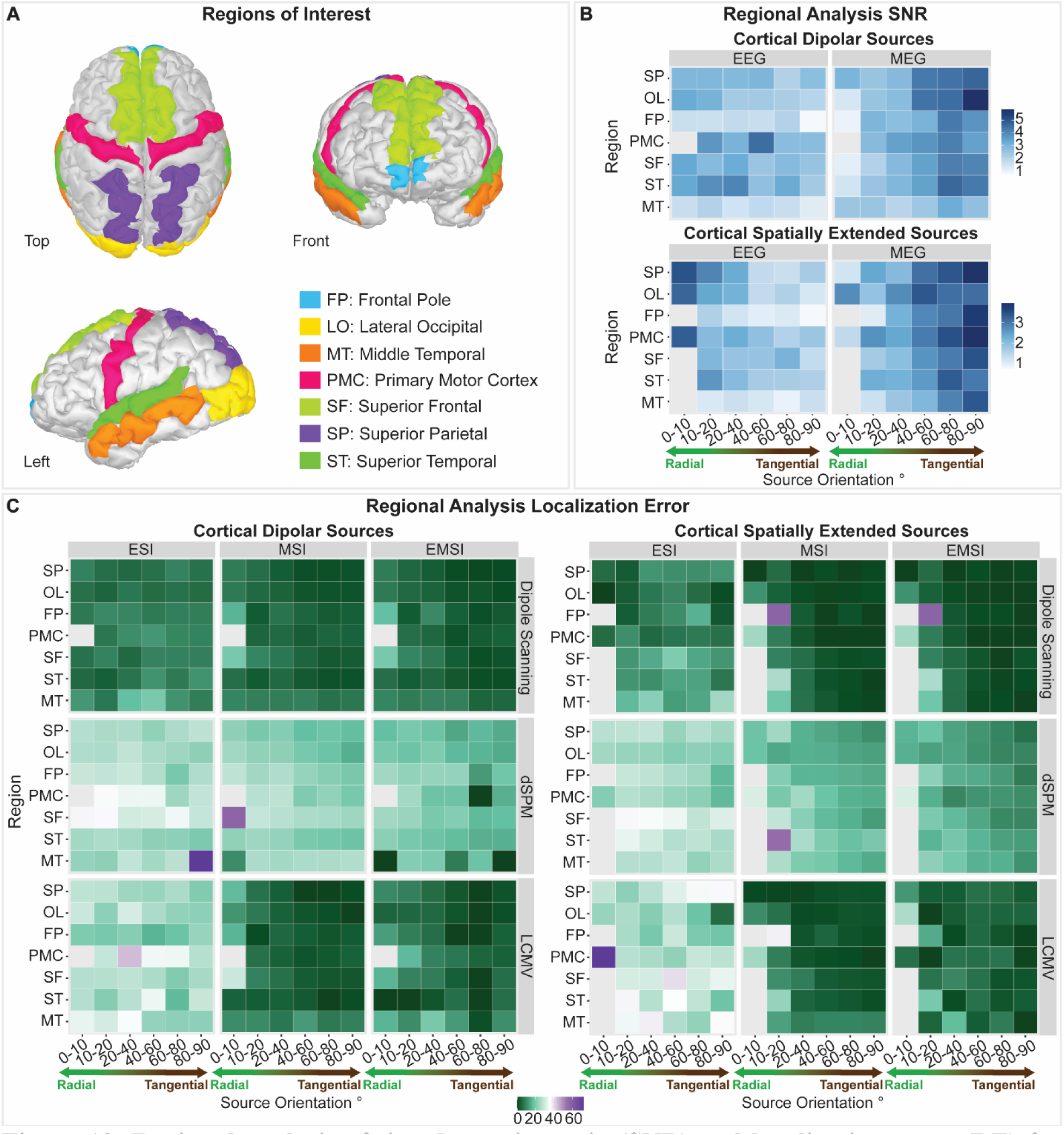
Regional analysis of signal-to-noise ratio (SNR) and localization error (LE) for both dipolar and spatially extended sources. (**A**) Seven epilepsy-relevant cortical regions included in the source space for both dipolar and spatially extended sources, with 24 simulations performed per region for each source type: Frontal Pole (FP), Lateral Occipital (LO), Middle Temporal (MT), Primary Motor Cortex (PMC), Superior Frontal (SF), Superior Parietal (SP), and Superior Temporal (ST). (**B**) Regional EEG and MEG SNR for both dipolar (top heatmaps) and spatially extended sources (bottom heatmaps) across all source orientations. (**C**) Regional electric source imaging (ESI), magnetic source imaging (MSI), and electromagnetic source imaging (EMSI) LE across inverse methods (dipole scanning, dSPM, and LCMV) and source orientations for dipolar (left heatmap block) and spatially extended (right heatmap block) sources. Median values are reported in all heatmap panels. Due to space limitations, asterisks indicating statistical significance are omitted.

Similar patterns were observed for spatially extended sources. EEG was more sensitive to radial sources located at the gyral crowns and sulcal bottoms, whereas MEG showed greater sensitivity to tangential sources along sulcal walls (**Fig. 12B**). Dipole scanning again yielded lower LE than both dSPM and LCMV (**Fig. 12C**), while MSI and EMSI showed greater localization accuracy for activity arising from sulcal walls (**Fig. 12C**). ESI showed no consistent orientation-dependent localization pattern (**Fig. 12C**). Across all seven cortical regions, MEG showed significantly higher SNR than EEG for spatially extended sources (*p*<0.001, Bonferroni corrected; **Fig. 12B**). Similarly, MSI and EMSI yielded significantly lower LE than ESI across all regions (*p*<0.001, Bonferroni corrected; **Fig. 12C**), with no significant differences between MSI and EMSI.

## 4. Discussion

In this study, we modeled realistic HD-EEG and MEG epileptic data using interictal iEEG recordings from patients with DRE and background noise segments from healthy controls to assess the sensitivity profiles of the two modalities and their localization accuracy in relation to the biophysical properties of epileptogenic generators. By varying source orientation, spatial extent, and location, we simulated 3,808 paired HD-EEG and MEG datasets spanning seven cortical regions (frontal pole, lateral occipital, middle temporal, primary motor cortex, superior frontal, superior parietal, and superior temporal), as well as the insular cortex and hippocampal complex. Both modalities showed a strong dependence on local source geometry, which was also reflected in localization performance. Four main findings support our conclusions: (i) EEG was more sensitive to radially oriented sources at gyral crowns and sulcal fundi, whereas MEG was more sensitive to tangentially oriented sources along sulcal walls; (ii) these sensitivities were complementary and largely non-overlapping, with MEG failing to detect 80–90% of radial sources and EEG failing to detect ∼80% of tangential sources (**Fig. 4B**); (iii) dipole scanning, despite its algorithmic simplicity, outperformed both dSPM and LCMV in overall localization accuracy across cortical dipolar, spatially extended, and insular sources; and (iv) MSI yielded the lowest LE for cortical and insular sources across inverse methods, while EMSI consistently outperformed ESI and performed comparably to MSI. Collectively, these findings provide evidence-based guidance for selecting non-invasive neuroimaging modalities and source localization methods to identify epileptogenic generators across anatomically and biophysically diverse brain regions during the presurgical evaluation of patients with DRE.

### 4.1. Anatomical preferences of EEG and MEG

Our sensitivity results extend prior simulation work [4,5,9,13,35,36,50] by enhancing ecological validity through the use of patient-derived spikes, physiological background noise from actual recordings, eight individual head models resulting in 3,808 simulations, and epileptologist-verified spike markings. We further modeled both dipolar and spatially extended generators, the former as a benchmark and the latter as a more realistic representation of synchronized neuronal activity. The orientation preferences described above were consistent across both source types, in line with prior reports showing that cortical anatomy modulates spike sensitivity in both modalities [4,5,36,51]. Notably, despite the general assumption that MEG is insensitive to radial generators [7], MEG detected a small proportion of radial-source spikes at attenuated SNR (dipolar sources: 11.45% at SNR: 4.46 ± 3.31; spatially extended sources: 4.94% at SNR: 1.74 ± 1.46), possibly reflecting the non-spherical geometry of the human head. For insular sources, neither modality showed a clear orientation-specific SNR pattern; however, EEG detected a higher proportion of spikes from sources with more radial orientations, whereas MEG detected more spikes from sources with more tangential orientations. For the hippocampal complex, both modalities detected only a small portion of low-SNR spikes, without clear modality-specific orientation preferences. These observations are consistent with previous studies suggesting that, as source depth increases, dipole orientation plays a progressively smaller role in spike detection [36]. Together, these anatomical preferences help explain why spikes are often visible in only one modality in clinical recordings [4,11–17]. In summary, MEG favors the detection of spikes generated by tangentially oriented sources without being entirely insensitive to radial and deep generators, whereas EEG preferentially detects epileptic activity arising from gyral crowns and sulcal bottoms.

### 4.2. Localization accuracy depends on source orientation and cortical geometry

Previous EEG and MEG sensitivity studies have focused primarily on SNR analyses of modeled epileptogenic sources [4,5,35,36,51], without examining how these sensitivity profiles relate to the localization performance of ESI, MSI, and EMSI. Having characterized spike detectability, we therefore evaluated localization accuracy using dipole scanning, the inverse method most widely used in clinical practice, alongside the more experimental dSPM and LCMV approaches.

For cortical dipolar, spatially extended, and insular sources, MSI showed a stronger dependence on source orientation than ESI and achieved its highest localization accuracy for tangential sources along the fissural cortex. ESI showed more stable localization performance across orientations but generally higher LE than MSI, particularly for sources with unfavorable anatomical profiles, such as those located deep within a sulcus. Consistent with this, MSI yielded lower LE than ESI across source orientations for dipolar, spatially extended, and insular generators, supporting the view that MEG can provide more focal estimates of epileptic activity than EEG [3].

Given the complementary sensitivity profiles of EEG and MEG, we next evaluated whether their integration through EMSI provided additional localization value. EMSI performance was largely driven by MSI: for sulcal wall sources, both MSI and EMSI localized tangential generators more accurately than ESI, and a similar advantage was observed for insular sources despite the absence of a clear orientation-dependent SNR pattern. These findings are consistent with reports showing that MEG is not entirely insensitive to deep sources and can provide additional diagnostic information for insular localization, complementing EEG findings even in patients with non-lesional MRI [33,52,53].

Across inverse methods, dipole scanning consistently yielded the lowest LE, outperforming both dSPM and LCMV in identifying the location of maximal epileptic activity. Because dipole scanning does not reconstruct source extent, we additionally evaluated dSPM and LCMV using AUC and SD. Both dSPM and LCMV methods showed limited ability to recover the spatial extent of the simulated generators, with EMSI-dSPM showing the most favorable combination of higher AUC and lower SD, consistent with previous findings [34,49]. These findings suggest that nonlinear distributed approaches, such as the maximum entropy on the mean (MEM), may be better suited for estimating the spatial extent of epileptogenic generators [34,37]. Overall, MEG and MSI detected and localized epileptic activity arising from sulcal walls more accurately than EEG and ESI, while dipole scanning provided the lowest LE among the inverse methods tested.

### 4.3. Limitations and future perspectives

A primary limitation of this study is that the proposed pipeline was evaluated exclusively on simulated data and has not yet been validated using real patient recordings. We also modeled only single-source activations with short-duration waveforms, leaving the sensor-level effects of simultaneously active generators unexplored. Although the use of patient-derived iEEG spikes and empirical background noise improved the realism of the simulations, using a single waveform for each source type assumes that discharges arising from gyral crowns, sulcal fundi, and lesions of different etiologies (e.g., FCD vs. hippocampal sclerosis) share similar temporal dynamics. This assumption does not capture the known electrophysiological heterogeneity of epileptogenic tissue. However, this choice was deliberate, as keeping the waveform constant within each source category (cortical, insular, and hippocampal) allowed direct comparisons of SNR and LE across source orientations and depths. Another consideration is that the differential sensitivity profiles of EEG and MEG only partly explain why some spikes are detected by a single modality. Noise level also plays an important role, as epileptic activity may be present in the other modality but remain undetected because of low SNR [4,54,55].

Future work should extend this framework in three dimensions: (1) evaluating nonlinear inverse approaches such as the MEM; (2) incorporating clinical recordings and simulations of multiple interacting sources to assess localization accuracy and spatial extent recovery under more realistic conditions; and (3) validating the pipeline using phantom studies, which provide controlled yet physically realistic representations of head geometry and recording conditions and could further characterize HD-EEG and MEG sensitivity across source configurations and pediatric pathologies [56,57].

## 5. Conclusions

We provide a comprehensive, ground-truth-based characterization of EEG and MEG sensitivity profiles and a direct comparison of ESI, MSI, and EMSI localization performance across a range of epileptogenic generator configurations. Both modalities exhibit distinct anatomical preferences that translate into differences in localization performance: ESI is more stable across orientations, whereas MSI and EMSI provide more focal and accurate localization for sources arising from sulcal walls. Clinically, these findings argue against treating HD-EEG and MEG as interchangeable or considering one modality merely confirmatory of the other. Because each modality may fail to detect sources preferentially captured by the other, simultaneous acquisition may provide important complementary information during the presurgical evaluation of patients with DRE, particularly when the suspected generator lies within the fissural cortex. EMSI largely preserved the localization advantage of MSI while outperforming ESI, suggesting that multimodal integration can combine the complementary sensitivities of EEG and MEG without sacrificing the strengths of either modality. This complementary information may improve non-invasive characterization of epileptogenic generators and help guide subsequent invasive evaluation and surgical planning. Ultimately, mapping each modality to its preferred anatomical targets may enable more tailored source imaging strategies for challenging lesion profiles such as bottom-of-sulcus dysplasia (BOSD), a highly epileptogenic form of FCD confined to the sulcal fundus [58].

## Acknowledgements

Funding for this research was provided by the U.S. National Institute of Neurological Disorders and Stroke (NINDS) under award no. R01NS104116 and R01NS134944 [Principal Investigator (PI): Christos Papadelis].

## Author contributions

**Glykeria Sdoukopoulou**: Writing – original draft, validation, methodology, investigation, formal analysis, data curation. **Saeed Jahromi**: Writing – review & editing. **Ludovica Corona**: Writing – review & editing. **Hmayag Partamian**: Writing – review & editing. **Jaehyung Lim**: Writing – review & editing. **Jawata Afnan**: Writing – review & editing, validation. **Christophe Grova**: Writing – review & editing, validation. **Christos Papadelis**: Writing – review & editing, validation, supervision, resources, methodology, investigation, formal analysis, data curation, conceptualization.

## Ethics statement

The protocol was approved by North Texas Regional Institutional Review Board (IRB-2019-166; PI: C. Papadelis; approval date: 06/24/2025).

## Data availability statement

The data used in this study will be available upon reasonable request.

## References

[1] C. Papadelis, M.S. Perry, Localizing the Epileptogenic Zone with Novel Biomarkers, Seminars in Pediatric Neurology 39 (2021) 100919. 10.1016/j.spen.2021.100919.

[2] M. Antonakakis, F. Kaiser, S. Rampp, S. Kovac, H. Wiendl, W. Stummer, J. Gross, C. Kellinghaus, M. Khaleghi-Ghadiri, G. Möddel, C.H. Wolters, Targeted and optimized multi-channel transcranial direct current stimulation for focal epilepsy: An N-of-1 trial., Brain Stimul 17 (2024) 221–223. 10.1016/j.brs.2024.02.010.

[3] D. Cohen, B.N. Cuffin, Demonstration of useful differences between magnetoencephalogram and electroencephalogram., Electroencephalogr Clin Neurophysiol 56 (1983) 38–51. 10.1016/0013-4694(83)90005-6.

[4] D.M. Goldenholz, S.P. Ahlfors, M.S. Hämäläinen, D. Sharon, M. Ishitobi, L.M. Vaina, S.M. Stufflebeam, Mapping the signal-to-noise-ratios of cortical sources in magnetoencephalography and electroencephalography, Human Brain Mapping 30 (2009) 1077–1086. 10.1002/hbm.20571.

[5] W.E. Kincses, C. Braun, S. Kaiser, T. Elbert, Modeling extended sources of event-related potentials using anatomical and physiological constraints, Hum. Brain Mapp. 8 (1999) 182–193. 10.1002/(SICI)1097-0193(1999)8:4%3C182::AID-HBM3%3E3.0.CO;2-M.

[6] S. Baillet, J.C. Mosher, R.M. Leahy, Electromagnetic brain mapping, IEEE Signal Process. Mag. 18 (2001) 14–30.

[7] M. Hämäläinen, R. Hari, R.J. Ilmoniemi, J. Knuutila, O.V. Lounasmaa, Magnetoencephalography—theory, instrumentation, and applications to noninvasive studies of the working human brain, Rev. Mod. Phys. 65 (1993) 413–497. 10.1103/RevModPhys.65.413.

[8] C.H. Wolters, A. Anwander, X. Tricoche, D. Weinstein, M.A. Koch, R.S. MacLeod, Influence of tissue conductivity anisotropy on EEG/MEG field and return current computation in a realistic head model: A simulation and visualization study using high-resolution finite element modeling, NeuroImage 30 (2006) 813–826. 10.1016/j.neuroimage.2005.10.014.

[9] S.P. Ahlfors, J. Han, J.W. Belliveau, M.S. Hämäläinen, Sensitivity of MEG and EEG to Source Orientation, Brain Topogr 23 (2010) 227–232. 10.1007/s10548-010-0154-x.

[10] E. Tamilia, M. AlHilani, N. Tanaka, M. Tsuboyama, J.M. Peters, P.E. Grant, J.R. Madsen, S.M. Stufflebeam., P.L. Pearl, C. Papadelis, Assessing the localization accuracy and clinical utility of electric and magnetic source imaging in children with epilepsy, Clinical Neurophysiology 130 (2019) 491–504. 10.1016/j.clinph.2019.01.009.

[11] M. Iwasaki, E. Pestana, R.C. Burgess, H.O. Lüders, H. Shamoto, N. Nakasato, Detection of Epileptiform Activity by Human Interpreters: Blinded Comparison between Electroencephalography and Magnetoencephalography, Epilepsia 46 (2005) 59–68. 10.1111/j.0013-9580.2005.21104.x.

[12] S. Knake, E. Halgren, H. Shiraishi, K. Hara, H.M. Hamer, P.E. Grant, V.A. Carr, D. Foxe, S. Camposano, E. Busa, T. Witzel, M.S. Hämäläinen, S.P. Ahlfors, E.B. Bromfield, P.M. Black, B.F. Bourgeois, A.J. Cole, G.R. Cosgrove, B.A. Dworetzky, J.R. Madsen, P.G. Larsson, D.L. Schomer, E.A. Thiele, A.M. Dale, B.R. Rosen, S.M. Stufflebeam, The value of multichannel MEG and EEG in the presurgical evaluation of 70 epilepsy patients, Epilepsy Research 69 (2006) 80–86. 10.1016/j.eplepsyres.2006.01.001.

[13] C. Baumgartner, E. Pataraia, G. Lindinger, L. Deecke, Neuromagnetic recordings in temporal lobe epilepsy., J Clin Neurophysiol 17 (2000) 177–189. 10.1097/00004691-200003000-00007.

[14] R. Hari, S. Baillet, G. Barnes, R. Burgess, N. Forss, J. Gross, M. Hämäläinen, O. Jensen, R. Kakigi, F. Mauguière, N. Nakasato, A. Puce, G.-L. Romani, A. Schnitzler, S. Taulu, IFCN-endorsed practical guidelines for clinical magnetoencephalography (MEG), Clinical Neurophysiology 129 (2018) 1720–1747. 10.1016/j.clinph.2018.03.042.

[15] Y.Y. Lin, Y.H. Shih, J.C. Hsieh, H.Y. Yu, C.H. Yiu, T.T. Wong, T.C. Yeh, S.Y. Kwan, L.T. Ho, D.J. Yen, Z.A. Wu, M.S. Chang, Magnetoencephalographic yield of interictal spikes in temporal lobe epilepsy, NeuroImage 19 (2003) 1115–1126. 10.1016/S1053-8119(03)00181-2.

[16] G. Ramantani, R. Boor, R. Paetau, N. Ille, R. Feneberg, A. Rupp, T. Boppel, M. Scherg, D. Rating, T. Bast, MEG Versus EEG: Influence of Background Activity on Interictal Spike Detection:, Journal of Clinical Neurophysiology 23 (2006) 498–508. 10.1097/01.wnp.0000240873.69759.cc.

[17] J.R. Stapleton-Kotloski, R.J. Kotloski, G. Popli, D.W. Godwin, Magnetoencephalography: Clinical and Research Practices, Brain Sciences 8 (2018) 157. 10.3390/brainsci8080157.

[18] H. Stefan, Magnetic brain source imaging of focal epileptic activity: a synopsis of 455 cases, Brain 126 (2003) 2396–2405. 10.1093/brain/awg239.

[19] I. Merlet, R. Paetau, L. García-Larrea, K. Uutela, M.-L. Granström, F. Mauguière, Apparent asynchrony between interictal electric and magnetic spikes:, NeuroReport 8 (1997) 1071–1076. 10.1097/00001756-199703240-00002.

[20] E. Rodin, M. Funke, P. Berg, F. Matsuo, Magnetoencephalographic spikes not detected by conventional electroencephalography, Clinical Neurophysiology 115 (2004) 2041–2047. 10.1016/j.clinph.2004.04.002.

[21] H.-M. Park, N. Nakasato, M. Iwasaki, H. Shamoto, T. Tominaga, T. Yoshimoto, Comparison of Magnetoencephalographic Spikes with and without Concurrent Electroencephalographic Spikes in Extratemporal Epilepsy, Tohoku J. Exp. Med. 203 (2004) 165–174. 10.1620/tjem.203.165.

[22] Y. Attal, B. Maess, A. Friederici, O. David, Head models and dynamic causal modeling of subcortical activity using magnetoencephalographic/electroencephalographic data, Revneuro 23 (2012) 85–95. 10.1515/rns.2011.056.

[23] Y. Attal, D. Schwartz, Assessment of Subcortical Source Localization Using Deep Brain Activity Imaging Model with Minimum Norm Operators: A MEG Study, PLoS ONE 8 (2013) e59856. 10.1371/journal.pone.0059856.

[24] W.W. Lytton, R. Orman, M. Stewart, Computer simulation of epilepsy: Implications for seizure spread and behavioral dysfunction, Epilepsy & Behavior 7 (2005) 336–344. 10.1016/j.yebeh.2005.06.011.

[25] F. Pizzo, N. Roehri, S. Medina Villalon, A. Trébuchon, S. Chen, S. Lagarde, R. Carron, M. Gavaret, B. Giusiano, A. McGonigal, F. Bartolomei, J.M. Badier, C.G. Bénar, Deep brain activities can be detected with magnetoencephalography, Nat Commun 10 (2019) 971. 10.1038/s41467-019-08665-5.

[26] S. Lee, S. Wu, J.X. Tao, S. Rose, P.C. Warnke, N.P. Issa, W. Van Drongelen, Manifestation of Hippocampal Interictal Discharges on Clinical Scalp EEG Recordings, Journal of Clinical Neurophysiology 40 (2023) 144–150. 10.1097/WNP.0000000000000867.

[27] P. Laoprasert, J.G. Ojemann, M.H. Handler, Insular epilepsy surgery, Epilepsia 58 (2017) 35–45. 10.1111/epi.13682.

[28] C. Abdallah, T. Hedrich, A. Koupparis, J. Afnan, J.A. Hall, J. Gotman, F. Dubeau, N. Von Ellenrieder, B. Frauscher, E. Kobayashi, C. Grova, Clinical Yield of Electromagnetic Source Imaging and Hemodynamic Responses in Epilepsy: Validation With Intracerebral Data, Neurology 98 (2022). 10.1212/WNL.0000000000200337.

[29] Ü. Aydin, S. Rampp, A. Wollbrink, H. Kugel, J.-H. Cho, T.R. Knösche, C. Grova, J. Wellmer, C.H. Wolters, Zoomed MRI Guided by Combined EEG/MEG Source Analysis: A Multimodal Approach for Optimizing Presurgical Epilepsy Work-up and its Application in a Multi-focal Epilepsy Patient Case Study, Brain Topography 30 (2017) 417–433. 10.1007/s10548-017-0568-9.

[30] R.K. Chikara, S. Jahromi, E. Tamilia, J.R. Madsen, S.M. Stufflebeam, P.L. Pearl, C. Papadelis, Electromagnetic source imaging predicts surgical outcome in children with focal cortical dysplasia, Clinical Neurophysiology 153 (2023) 88–101. 10.1016/j.clinph.2023.06.015.

[31] R.A. Chowdhury, I. Merlet, G. Birot, E. Kobayashi, A. Nica, A. Biraben, F. Wendling, J.M. Lina, L. Albera, C. Grova, Complex patterns of spatially extended generators of epileptic activity: Comparison of source localization methods cMEM and 4-ExSo-MUSIC on high resolution EEG and MEG data, NeuroImage 143 (2016) 175–195. 10.1016/j.neuroimage.2016.08.044.

[32] L. Corona, S. Rijal, O. Tanritanir, S. Shahdadian, C.G. Keator, L. Tran, S.I. Malik, M. Bosemani, D. Hansen, D. Shahani, M.S. Perry, C. Papadelis, Electromagnetic Source Imaging in Presurgical Evaluation of Children with Drug-Resistant Epilepsy, JoVE (2024) e66494. 10.3791/66494.

[33] C. Papadelis, S.E. Conrad, Y. Song, S. Shandley, D. Hansen, M. Bosemani, S. Malik, C. Keator, M.S. Perry, Case Report: Laser Ablation Guided by State of the Art Source Imaging Ends an Adolescent’s 16-Year Quest for Seizure Freedom, Front. Hum. Neurosci. 16 (2022) 826139. 10.3389/fnhum.2022.826139.

[34] R.A. Chowdhury, J.M. Lina, E. Kobayashi, C. Grova, MEG Source Localization of Spatially Extended Generators of Epileptic Activity: Comparing Entropic and Hierarchical Bayesian Approaches, PLoS ONE 8 (2013) e55969. 10.1371/journal.pone.0055969.

[35] M.C. Piastra, A. Nüßing, J. Vorwerk, M. Clerc, C. Engwer, C.H. Wolters, A comprehensive study on electroencephalography and magnetoencephalography sensitivity to cortical and subcortical sources, Human Brain Mapping 42 (2021) 978–992. 10.1002/hbm.25272.

[36] A. Hunold, M.E. Funke, R. Eichardt, M. Stenroos, J. Haueisen, EEG and MEG: sensitivity to epileptic spike activity as function of source orientation and depth, Physiol. Meas. 37 (2016) 1146–1162. 10.1088/0967-3334/37/7/1146.

[37] J. Afnan, Z. Cai, J. Lina, C. Abdallah, E. Delaire, T. Avigdor, V. Ros, T. Hedrich, N. Von Ellenrieder, E. Kobayashi, B. Frauscher, J. Gotman, C. Grova, EEG/MEG source imaging of deep brain activity within the maximum entropy on the mean framework: Simulations and validation in epilepsy, Human Brain Mapping 45 (2024) e26720. 10.1002/hbm.26720.

[38] Y. Attal, M. Bhattacharjee, J. Yelnik, B. Cottereau, J. Lefevre, Y. Okada, E. Bardinet, M. Chupin, S. Baillet, Modeling and Detecting Deep Brain Activity with MEG & EEG, in: 2007 29th Annual International Conference of the IEEE Engineering in Medicine and Biology Society, IEEE, Lyon, France, 2007: pp. 4937–4940. 10.1109/IEMBS.2007.4353448.

[39] B. Fischl, FreeSurfer, NeuroImage 62 (2012) 774–781. 10.1016/j.neuroimage.2012.01.021.

[40] E.T. Rolls, C.-C. Huang, C.-P. Lin, J. Feng, M. Joliot, Automated anatomical labelling atlas 3, NeuroImage 206 (2020) 116189. 10.1016/j.neuroimage.2019.116189.

[41] F. Tadel, S. Baillet, J.C. Mosher, D. Pantazis, R.M. Leahy, Brainstorm: A User-Friendly Application for MEG/EEG Analysis, Computational Intelligence and Neuroscience 2011 (2011) 1–13. 10.1155/2011/879716.

[42] A. Gramfort, T. Papadopoulo, E. Olivi, M. Clerc, Forward Field Computation with OpenMEEG, Computational Intelligence and Neuroscience 2011 (2011) 1–13. 10.1155/2011/923703.

[43] S. Lloyd, Least squares quantization in PCM, IEEE Transactions on Information Theory 28 (1982) 129–137. 10.1109/TIT.1982.1056489.

[44] S. Murakami, Y. Okada, Contributions of principal neocortical neurons to magnetoencephalography and electroencephalography signals, The Journal of Physiology 575 (2006) 925–936. 10.1113/jphysiol.2006.105379.

[45] L. Corona, E. Tamilia, M.S. Perry, J.R. Madsen, J. Bolton, S.S.D. Stone, S.M. Stufflebeam, P.L. Pearl, C. Papadelis, Non-invasive mapping of epileptogenic networks predicts surgical outcome, Brain 146 (2023) 1916–1931. 10.1093/brain/awac477.

[46] M.A.G. Matarrese, A. Loppini, L. Fabbri, E. Tamilia, M.S. Perry, J.R. Madsen, J. Bolton, S.S.D. Stone, P.L. Pearl, S. Filippi, C. Papadelis, Spike propagation mapping reveals effective connectivity and predicts surgical outcome in epilepsy, Brain 146 (2023) 3898–3912. 10.1093/brain/awad118.

[47] A.M. Dale, A.K. Liu, B.R. Fischl, R.L. Buckner, J.W. Belliveau, J.D. Lewine, E. Halgren, Dynamic Statistical Parametric Mapping: Combining fMRI and MEG for High-Resolution Imaging of Cortical Activity, Neuron 26 (2000) 55–67. 10.1016/S0896-6273(00)81138-1.

[48] B.D. Van Veen, W. Van Drongelen, M. Yuchtman, A. Suzuki, Localization of brain electrical activity via linearly constrained minimum variance spatial filtering, IEEE Trans. Biomed. Eng. 44 (1997) 867–880. 10.1109/10.623056.

[49] C. Grova, J. Daunizeau, J.-M. Lina, C.G. Bénar, H. Benali, J. Gotman, Evaluation of EEG localization methods using realistic simulations of interictal spikes, NeuroImage 29 (2006) 734–753. 10.1016/j.neuroimage.2005.08.053.

[50] S.P. Ahlfors, J. Han, F. Lin, T. Witzel, J.W. Belliveau, M.S. Hämäläinen, E. Halgren, Cancellation of EEG and MEG signals generated by extended and distributed sources, Human Brain Mapping 31 (2010) 140–149. 10.1002/hbm.20851.

[51] A. Hillebrand, G.R. Barnes, A Quantitative Assessment of the Sensitivity of Whole-Head MEG to Activity in the Adult Human Cortex, NeuroImage 16 (2002) 638–650. 10.1006/nimg.2002.1102.

[52] C. Solanki, J. Williams, C. Andrews, I. Fayed, C. Wu, Insula in epilepsy – “untying the gordian knot”: A systematic review, Seizure: European Journal of Epilepsy 106 (2023) 148–161. 10.1016/j.seizure.2023.02.019.

[53] T. Yu, D. Ni, X. Zhang, X. Wang, L. Qiao, X. Zhou, Y. Wang, Y. Li, G. Zhang, The role of magnetoencephalography in the presurgical evaluation of patients with MRI-negative operculo-insular epilepsy., Seizure 61 (2018) 104–110. 10.1016/j.seizure.2018.07.005.

[54] H. Yoshinaga, T. Nakahori, Y. Ohtsuka, E. Oka, Y. Kitamura, H. Kiriyama, K. Kinugasa, K. Miyamoto, T. Hoshida, Benefit of simultaneous recording of EEG and MEG in dipole localization., Epilepsia 43 (2002) 924–928. 10.1046/j.1528-1157.2002.42901.x.

[55] M. Zijlmans, G.M. Huiskamp, F.S.S. Leijten, W.M. Van Der Meij, G. Wieneke, A.C. Van Huffelen, Modality-specific spike identification in simultaneous magnetoencephalography/electroencephalography: a methodological approach., J Clin Neurophysiol 19 (2002) 183–191. 10.1097/00004691-200206000-00001.

[56] S. Jahromi, G. Sdoukopoulou, R.K. Chikara, S.M. Stufflebeam, M.P. Ottensmeyer, G. De Novi, C. Papadelis, 3D printed pediatric head phantom for assessing deep epileptic sources localization, Computers in Biology and Medicine 202 (2026) 111449. 10.1016/j.compbiomed.2026.111449.

[57] C. Papadelis, V. Poghosyan, P.B.C. Fenwick, A.A. Ioannides, MEG’s ability to localise accurately weak transient neural sources, Clinical Neurophysiology 120 (2009) 1958–1970. 10.1016/j.clinph.2009.08.018.

[58] E. Macdonald-Laurs, A.E.L. Warren, P. Francis, S.A. Mandelstam, W.S. Lee, M. Coleman, S.E.M. Stephenson, S. Barton, C. D’Arcy, P.J. Lockhart, R.J. Leventer, A.S. Harvey, The clinical, imaging, pathological and genetic landscape of bottom-of-sulcus dysplasia, Brain 147 (2024) 1264–1277. 10.1093/brain/awad379.

